# cAMP prevents antibody-mediated thrombus formation in COVID-19

**DOI:** 10.1101/2020.12.15.20247775

**Authors:** Jan Zlamal, Karina Althaus, Hisham Jaffal, Lisann Pelzl, Anurag Singh, Andreas Witzemann, Helene Häberle, Valbona Mirakaj, Peter Rosenberger, Tamam Bakchoul

## Abstract

Thromboembolic events are frequently reported in patients infected with the SARS-CoV-2 virus. However, the exact mechanisms of thromboembolic events remain elusive. In this work, we show that immunoglobulin G (IgG) subclass in patients with COVID-19 trigger the formation of procoagulant PLTs in a Fc-gamma-RIIA (FcγRIIA) dependent pathway leading to increased thrombus formation in vitro. Most importantly, these events were significantly inhibited via FcγRIIA blockade as well as by the elevation of PLTs’ intracellular cyclic-adenosine-monophosphate (cAMP) levels by the clinical used agent Iloprost. The novel findings of FcγRIIA mediated prothrombotic conditions in terms of procoagulant PLTs leading to higher thrombus formation as well as the successful inhibition of these events via Iloprost could be promising for the future treatment of the complex coagulopathy observed in COVID-19 disease.

**Key points:**

- Fc-gamma-receptor IIA mediated PS externalization on the PLT surface triggers increased thrombus formation
- Inductors of cAMP inhibit antibody-mediated thrombus formation and may have potential therapeutic advantage in COVID-19

## Introduction

Infection with SARS-CoV-2 has been shown to be associated with abnormalities in the coagulation system with an increased incidence of thromboembolic events in small vessels leading to higher mortality (1-3). Accumulating evidence suggests upregulated release of inflammatory cytokines and increased interaction between different actors of innate and adaptive immunity as the main causes for the prothrombotic environment observed in COVID-19 disease (4). Moreover, a significant number of reports described platelet (PLT) hyperactivity in patients with COVID-19, which is assumed to contribute to prothrombotic conditions (5, 6). Procoagulant PLTs, predominantly generated at the outer side of the growing thrombus, are increasingly recognized to link primary with secondary haemostasis (7-10). The latter is mainly executed by negatively charged membrane phospholipids externalized on the PLT surface. This unique feature of procoagulant PLTs enables the assembly of tenase as well as prothrombinase complexes leading to high thrombin burst, increased fibrin deposition and thrombus formation (11). Recently, we showed that PLTs from patients with severe COVID-19 infection express a procoagulant phenotype. Immunoglobulin G (IgG) fractions were found to be responsible for the COVID-19 induced procoagulant PLTs (Althaus et al. accepted).

In the current study, we investigate the time course of the generation of antibody-induced procoagulant PLTs as well as the mechanisms leading to alterations in the PLT phenotype in COVID-19. We observed that IgG fractions from severe COVID-19 patients induce increased thrombus formation in an Fc-gamma RIIA (FcγRIIA) dependent manner. More importantly, we were able to show that cyclic-adenosine-monophosphate (cAMP) elevation in PLTs prevents COVID-19 antibody-induced procoagulant PLT generation as well as clot formation.

## Materials and Methods

### Patients and sera

Experiments were performed using leftover serum material from COVID-19 patients who were referred to our laboratory between March and June 2020. The diagnosis of SARS-CoV-2 infection was confirmed by real time PCR on material collected by nasal swabs. Sera from ICU non-Covid-19 patients with postoperative sepsis were collected to serve as ICU control group. Additionally, sera were collected from healthy blood donors at the Blood Donation Centre Tuebingen after written consensus was obtained to establish cutoff values when appropriate. Serum samples were stored at −80°C and thawed at room temperature prior to the performed experimental procedure. All sera were heat-inactivated at 56°C for 30 minutes (min), which was followed by a centrifugation step at 5000xg. The spun-down was discarded and supernatants were handled as described in the following sections.

### IgG preparation

IgG fractions were isolated by the use of a commercially available IgG-purification-kit (Melon™-Gel IgG Spin Purification Kit, Thermo Fisher Scientific, Waltham, USA) and used as recommended by the manufacturer. In brief, heat inactivated serum was diluted 1:10 in purification buffer and incubated with the kit specific Gel IgG Purification Support over four cycles for 10 min. Subsequently, periodically performed centrifugation steps through a 10 µm pore size filter tube were performed for 1 min at 5000xg. The flow throw was collected into 100 kDa-pore sized centrifugal filters (Amicon Ultra-4, Merck Millipore, Cork, Ireland) with subsequent concentration to the initial volume of the used serum sample via centrifugation (10-15 min, 2000xg, 4°C, with brake). Afterwards, IgG concentrations were measured at a mass extinction coefficient of 13.7 at 280 nm wavelength using a NanoDrop One spectrophotometer (VWR, Bruchsal, Germany). IgG purity was verified using Coomassie staining (Abcam, Cambridge, UK).

### Preparation of washed platelets

Washed platelets (wPLTs) were prepared from venous blood samples as described previously (12). Briefly, whole blood from healthy donors was withdrawn by cubital venipuncture into acidic-dextrose containing vacutainers (Becton-Dickinson, Plymouth, UK) and allowed to rest for 45 min at 37°C. After centrifugation (20 min, 120xg, room temperature [RT], no brake) PLT-rich-plasma (PRP) was gently separated and supplemented with apyrase (5 µL/mL, Sigma-Aldrich, St. Louis, USA) and prewarmed ACD-A (111 µL/mL, Terumo BCT, Inc., Lakewood, USA). After an additional centrifugation step (7 min, 650xg, RT, no brake), the PLT pellet was resuspended in 5 mL of wash-solution (modified Tyrode buffer: 5 mL bicarbonate buffer, 20 percent (%) bovine serum albumin, 10% glucose solution [Braun, Melsungen, Germany], 2.5 U/mL apyrase, 1 U/mL hirudin [Pentapharm, Basel, Swiss], pH 6.3) and allowed to rest for 15 min at 37°C. Following final centrifugation (7 min, 650xg, RT, no brake) wPLTs were resuspended in 2 mL of resuspension-buffer (50 mL of modified Tyrode buffer, 0.5 mL of 0.1 M MgCl2, 1 mL of 0.2 M CaCl2, pH 7.2) and adjusted to 300×109 PLTs/L after the measurement at a Cell-Dyn Ruby hematological analyzer (Abott, Wiesbaden, Germany) was performed. For calcium chelation experiments, the PLT pellet was resuspended with resuspension-buffer without the supplementation of calcium (50 mL of modified Tyrode buffer, 0.5 mL of 0.1 M MgCl2, pH 7.2).

### Treatment of wPLTs with ICU COVID-19 sera/IgG

wPLTs (7.5×106) were supplemented with 5 µL serum/IgG from ICU COVID-19 patients or control serum/IgG and incubated for 1.5 hours (hs) at RT under rotating conditions. Afterwards, samples were washed once (7 min, 650xg, RT, no brake), resuspended in 100 µL of phosphate buffered saline (PBS, [Biochrom, Berlin, Germany]) and further handled as described in the following sections.

### Detection of phosphatidylserine exposure

To assess externalization of phosphatidylserine (PS) on the PLT surface after antibody treatment, 10 µL of PLT suspension were transferred into 100 µL of Hank’s balanced salt solution (HBSS), (137 mM NaCl, 1.25 mM CaCl2, 5.5 mM glucose, [Carl-Roth, Karlsruhe, Germany]) and incubated with 1 µL Annexin V-FITC (Immunotools, Friesoythe, Germany) for 30 min at RT in the dark. To induce the maximal externalization of PS on the PLT surface, wPLTs were incubated with ionomycin (Sigma-Aldrich, St. Louis, USA, [5 µM, 15 min at RT]). Afterwards, PLTs were filled up with HBSS to a final volume of 500 µL and acquired at a flow cytometer ([FC], Navios, Beckman-Coulter, Brea, USA).

PLTs were gated based on their characteristic forward scatter (FSC) vs. side scatter (SSC) properties as well as CD41a and CD42a expression (anti-CD41a-PC5 and anti-CD42a-PerCP, both BD, San Jose, USA), respectively. Test results were analyzed as fold increase of the percentage PS positive PLTs compared to PLTs that were incubated with serum/IgG from healthy individuals.

### Determination of changes in the inner-mitochondrial-transmembrane potential (ΔΨ)

Changes of the mitochondrial inner transmembrane potential (ΔΨ) induced by ICU COVID-19 sera/IgG were analyzed by FC, as previously reported (13). Briefly, after treatment with IgG/serum from ICU COVID-19 patients, wPLTs (∼2×10^6^) were incubated with a final concentration of 10 µM tetramethylrhodamine, ethyl ester (TMRE, Abcam, Cambridge, UK) for 30 min at RT in the dark. Carbonyl cyanide 4-(trifluoromethoxy) phenylhydrazone (FCCP, Abcam, Cambridge, UK) which is an uncoupler of mitochondrial oxidative phosphorylation served as positive control in each experiment. After staining with TMRE, PLTs were filled up with PBS to a final volume of 500 µL and immediately analyzed by FC. Changes in the ΔΨ were determined in gated PLTs as percentage of TMRE negative events and normalized to PLTs that were treated with serum/IgG from healthy controls.

### Phenotyping of different PLT-populations

ICU COVID-19 serum/IgG-mediated procoagulant changes (PS/CD62p-double positive) were assessed in some experiments using a triple staining. Briefly, 10 µL (∼1×10^6^) of the resuspended PLT suspension were transferred into 10 µL of HBSS and incubated with 1 µL anti-CD62p-APC (BD, San Jose, USA), 1 µL Annexin-FITC (Immunotools, Friesoythe, Germany) and 1 µL anti-CD42a-PerCP (BD, San Jose, USA) for 15 min at RT in the dark. PLTs treated with thrombin receptor activating peptide (TRAP-6, [20 µM, 30 min at RT]) and ionomycin (5 µM, 15 min at RT, [both Sigma-Aldrich, St. Louis, USA]) served as positive controls. Afterwards, PLTs were resuspended with HBSS to a final volume of 500 µL and immediately assessed via FC. In selected experiments that were designed to investigate the impact of calcium on cell signalling, PLT PS externalization was assessed by the use of the calcium independent marker lactadherin. 1 µL of lactadherin-FITC (Haematologic Technologies, Essex Junction, USA) was incubated with wPLTs as for Annexin-FITC for 15 min at RT, in the dark, and the PLT suspension filled up to final volume of 500 µL with PBS prior to FC analysis.

### Western blot analysis of caspase 3 cleavage

Protein levels of cleaved-caspase 3 were determined by western blot. After serum/IgG incubation, cells were washed with PBS for 7 min, 700xg at 4°C. Subsequently, the pellet was resuspended in 100 μL of ice-cold RIPA lysis buffer containing HALT™ protease and phosphatase inhibitor-cocktail (both ThermoFisher Scientific, Paisley, UK). Protein concentrations were determined using the NanoDrop One spectrophotometer (VWR, Bruchsal, Germany). 100 µg of protein was solubilized in sample buffer (Invitrogen™, Carlsbad, USA) at 95°C for 10 min. Proteins were separated by electrophoresis using 12% SDS-PAGE gels in glycine-tris buffer. Thereafter, probes were transferred to polyvinylidene difluoride (PVDF) membranes (0.45 µm, Merck, Tullagreen, Ireland). Afterwards, membranes were blocked with 5% milk in tris-buffered saline (TBS-T, 20 mM Tris, 140 mM NaCl, 0.1% Tween, pH 7.6) at RT for 1 h. Membranes were then incubated with primary anti-human cleaved-caspase 3 antibody (1:1000, Abcam, Cambridge, UK) and anti-human alpha-tubulin (1:1000, Cell Signaling Technology, Danvers, USA) at 4°C overnight. After washing with TBS-T buffer, the membranes were incubated with the appropriate secondary anti-rabbit or anti-mouse antibody conjugated with IRDye®680 / IRDye®800 (1:3000, LI-COR®, Lincoln, USA) for 1 h at RT. Protein bands were detected after additional washes (TBS-T) with Odyssey infrared imaging system (LI-COR®, Lincoln, USA). Western blots were analyzed by ImageJ software (NIH, Bethesda, USA). The results are shown as the ratio of total cleaved-caspase 3 to procaspase 3 (full fragment) and normalized to wPLTs that were treated with healthy control serum/IgG.

### Assessment of the mechanisms of antibody-mediated effects on PLTs

75 µL wPLTs were pretreated with the FcγRIIA blocking monoclonal antibody (moAb) anti-CD32 (5 µL moAb IV.3; stemcell™ technologies, Vancouver, Canada) for 45 min at 37°C prior to serum/IgG treatment. A monoclonal isotype (moAb) served as vehicle control ([SC-2025], Santa Cruz Biotechnology, Dallas, USA).

For the chelation of extracellular calcium, the non-membrane permeable chelator of extracellular calcium EGTA (Ethyleneglycol-bis(2-aminoethylether)-N,N,N′,N′-tetraacetic acid, 1 mM, 5 min at 37°C [Sigma Aldrich, St. Louis, USA]) was used.

For the depletion of calcium in the inner compartment of PLTs, the intracellular chelator of calcium BAPTA-AM (1,2-Bis(2-aminophenoxy)ethane-N,N,N’,N’-tetraacetic acid tetrakis(acetoxymethyl) ester, 20 µM, 15 min at 37°C, [Selleck, Houston, USA]) was used.

To investigate the effect of increased intracellular levels of cAMP, wPLTs were pretreated with the adenylate cyclase (ADC) inducers Forskolin (2.25 µM) and Iloprost (20 nM, both Sigma-Aldrich, St. Louis, USA) prior to the incubation with sera/IgGs from ICU COVID-19 patients.

### Investigations of antibody-mediated thrombus formation

To assess the impact of ICU COVID-19 IgG-induced effects on clot formation, an ex vivo model for thrombus formation was established. A microfluidic system (BioFlux, Fluxion Biosciences, Alameda, USA) was used at a shear rate of 1500^-1^ (60 dyne) according to the recommendations of the ISTH standardization committee for biorheology (14). Briefly, microfluidic channels were coated with collagen (100 µg/mL, Collagen Horm, Takeda, Linz, Austria) overnight at 4°C and blocked with 2.5% of human serum albumin (HSA, Kedrion, Barga, Italy) 1 h before perfusion at RT.

Whole blood samples of healthy individuals of blood group O were collected into hirudin containing monovettes (Sarstedt, Nuembrecht, Germany) and allowed to rest for 30 min at RT. After splitting the whole blood into aliquots of 200 µL, PRP was prepared via centrifugation (20 min, 120xg, at RT, no break). Afterwards, 45 µL of the supernatant PRP was gently separated and incubated with 5 µL of control or ICU COVID-19 IgG fractions and incubated for 90 min at RT under rotating conditions. 15 min prior to the end of the incubation period, Calcein-FITC (4 µM, [Thermo Scientific, Eugene, Oregon, USA]) was added to each sample. Subsequently, PRP was gently added back to reconstitute whole blood samples.

When indicated, the separated PRP was pretreated with moAb IV.3 or moAb isotype control at a concentration of 20 µg/mL for 30 min at RT. For cAMP induction, PRP was pretreated with Iloprost (20 nM) or vehicle control (5 min, 37°C) prior to IgG incubation.

Finally, reconstituted whole blood samples were run at a shear rate of 1500s^-1^ (60 dyne) for a maximum of 5 min. Immunofluorescence and bright field images were taken from 3-5 randomly chosen microscopic fields (x20, Olympus IX73, Olympus GmbH, Hamburg, Germany). Clot formation was assessed by measuring the % of surface area coated by thrombus (SAC) of 3-5 images via ImageJ (NIH, Bethesda, USA) and normalized to the whole area.

### Statistical analysis

Statistical analyses were performed using GraphPad Prism 7 (La Jolla, USA). T-test was used to analyze normally distributed results. Non-parametric test (Mann-Whitney test) was used when data failed to follow a normal distribution as assessed by D’Agostino and Pearson omnibus normality test. Group comparison was performed using the Wilcoxon rank-sum test and the Fisher exact test with categorical variables. A p-value <0.05 was assumed to represent statistical significance.

### Ethics

Studies involving human material were approved by the Ethics Committee of the Medical Faculty, Eberhard-Karls University of Tuebingen, Germany, and were conducted in accordance with the declaration of Helsinki.

## Results

### Patient characteristics

Sera from 26 ICU COVID-19 patients were enrolled in this study between March the 1^st^ and June the 16^th^ 2020. 21 of these ICU COVID-19 patients were included in a previous study (Althaus et al. accepted). The mean age of ICU COVID-19 patients was 58 years (range: 29-88 years). 20/30 (67%) patients had known risk factors for severe COVID-19 infection as described previously (15), including hypertension 18/30 (60%), obesity 6/30 (20%), coronary artery disease 4/30 (13%) and diabetes mellitus 6/30 (20%). Elevated D-Dimer levels were detected in all patients (median, range: 3.4 mg/dL, 0.9-45.0 mg/dL) and thrombosis was diagnosed in 13/30 (43%) patients. Longitudinal blood samples were available from four COVID-19 patients who were first admitted to normal ward and later to the ICU for mechanical ventilation. As ICU control group, 5 patients who were admitted to ICU due to non-COVID-19 related causes were included in this study.

### Sera from ICU COVID-19 patients induce progressive increase of procoagulant PLTs

To investigate whether sera of ICU COVID-19 patients have the potential to induce an increased Δψ depolarization as well as PS externalization on the PLT surface, wPLTs from healthy individuals were incubated with sera from 26 ICU COVID-19 patients with a severe course of disease as well as 5 ICU non-COVID-19 patients. Based on the calculated cuttoffs (mean+2xSD of healthy controls), 19/26 (73%) sera from patients with severe COVID-19 disease induced significantly higher Δψ depolarization in PLTs from healthy donors compared to ICU controls (FI in % Δψ depolarization±SEM: 6.10±1.12 vs. 0.67±0.10, p value <0.0001, Fig. 1 A). In addition, significantly higher PS externalization was observed when ICU COVID-19 sera were incubated with PLTs compared to ICU control sera (FI in % PS±SEM: 2.12±0.19 vs. 1.12±0.08, p value <0.0001, Fig. 1 B).

**Fig. 1.**
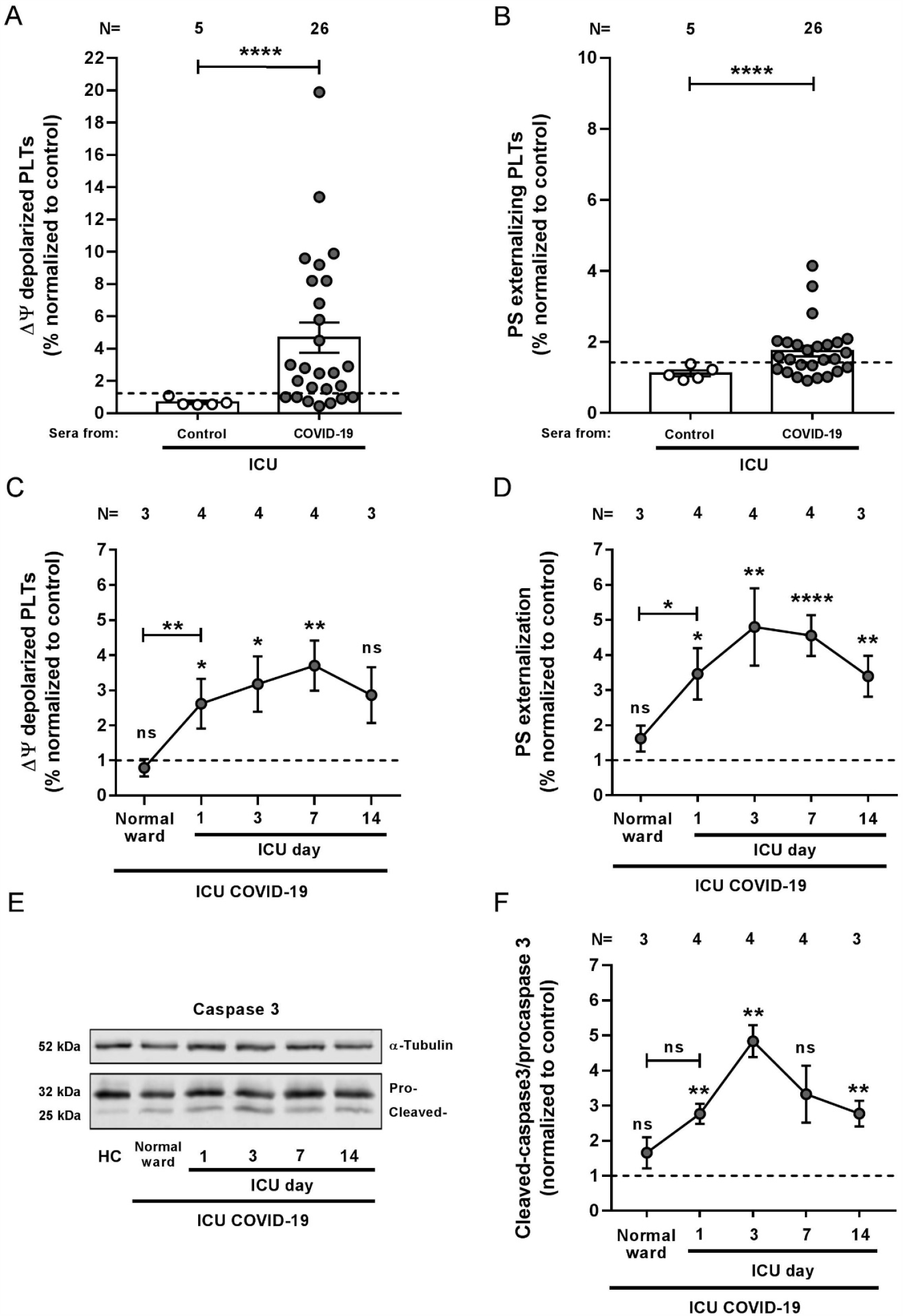
ICU COVID-19 patient serum induced effects on PLTs during disease. (**A+B**) ICU COVID-19 (n=26) or ICU non-COVID-19 control (n=5) patient serum induced changes in Δψ as well as PS externalization in wPLTs were analyzed by FC. (**C+D**) Sera of 4 ICU COVID-19 patients were collected for up to 14 days during hospitalization and analyzed for their ability to induce changes in Δψ as well as PS externalization in wPLTs via FC. (**E**) Representative WB image of detected cleaved caspase 3 (cleaved-) and procaspase 3 (pro-) levels in wPLTs that were incubated with follow up sera of one ICU COVID-19 patient (indicated in **C+D**). (**F**) Densitometric analysis of cleaved caspase 3/procaspase 3 ratios in wPLTs that were incubated with follow up sera of ICU COVID-19 patients (n=4, [indicated in **C+D**]) normalized to control. α-Tubulin served as loading control. Data are presented as mean±SEM of the measured fold increase compared to control. ns, not significant; *p<0.05, **p<0.01, ***p<0.001 and ****p<0.0001. The number of patient sera tested is reported in each graph. Dot lines in (**A+B**) represent the calculated cutoffs determined testing sera from healthy donors as mean of fold increase (FI)+2xSEM. HC, healthy control; Δψ, inner mitochondrial transmembrane potential; N, number of HCs or patients; PS, phosphatidylserine.

Next, we sought to investigate the time course of the observed changes in both markers. Longitudinal blood samples were available from four ICU COVID-19 patients. Sera were collected at hospital admission (normal ward or ICU) and during a follow up period at ICU for up to 14 days. As shown in Fig. 1 C-F, sera from ICU COVID-19 patients induced significant changes in Δψ depolarization, PS externalization and caspase 3 cleavage as clinical manifestation worsen requiring admission to ICU. Of note, antibody-induced changes peaked within day 3 and 7 of the ICU stay (FI in % Δψ depolarization±SEM: 3.71±0.72, p value 0.0070; and % PS externalization ±SEM: 4.80±1.11, p value <0.0001, respectively, Suppl. Fig. 1). These findings were further supported by WB analyses, as PLTs incubated with the corresponding ICU COVID-19 serum induced caspase 3 cleavage in similar kinetic (Ratio of cleaved caspase 3/procaspase 3 normalized to control±SEM: 4.84±0.45, p value 0.0035, Fig. 1 E+F). Notably, the rise of PLT markers was associated with increasing levels of detected IgGs against the spike S protein of SARS-CoV-2 in the corresponding ICU COVID-19 patients’ follow up sera but not in the total IgG contents of isolated IgG fractions (Suppl. Fig. 2 A and B, respectively). Moreover, declining PLT-counts were observed as Δψ depolarization as well as PS externalization increased, vice versa (Suppl. Fig. 3 A and B, respectively).

### IgGs from severe COVID-19 trigger procoagulant PLTs with increased ability to form blood clots

To further verify the impact of sera from severe COVID-19 patients on PLTs, the expression of the alpha granule release and PLT activation marker CD62p was analyzed in two colour FC in parallel to PS. FC analyses revealed that IgG fractions from severe COVID-19 patients induce remarkable changes in the PLT SSC and FSC (Fig. 2.1. A+C), as well as in the distribution of CD62p/PS positivity (Fig. 2.1. B+D). In contrast, the PLT population was almost non-affected after incubation with IgGs from healthy controls (HCs) or ICU non-COVID-19 control patients. Overall, higher percentage of double positive events was observed after incubation with IgGs from ICU COVID-19 patients compared to ICU and to HCs (% CD62p/PS positive PLTs±SEM: 31.63±3.86 vs. 4.04±1.16, p value 0.0007; and vs. 2.88±0.52, p value <0.0001, respectively, Fig. 2.1. II.). Additionally, significant elevation was observed in the PS single positive PLT population (% PS positive PLTs±SEM: 13.17±2.05 vs. 2.66±0.52, p value <0.0001; and vs. 2.36±0.29, p value 0.0002, Fig. 2.1. I) but not in the percentage of CD62p single positive PLTs (% CD62p positive PLTs±SEM: 19.40±1.83 vs. 13.53±1.90, p value 0.1011, Fig. 2.1. III).

**Fig. 2.**
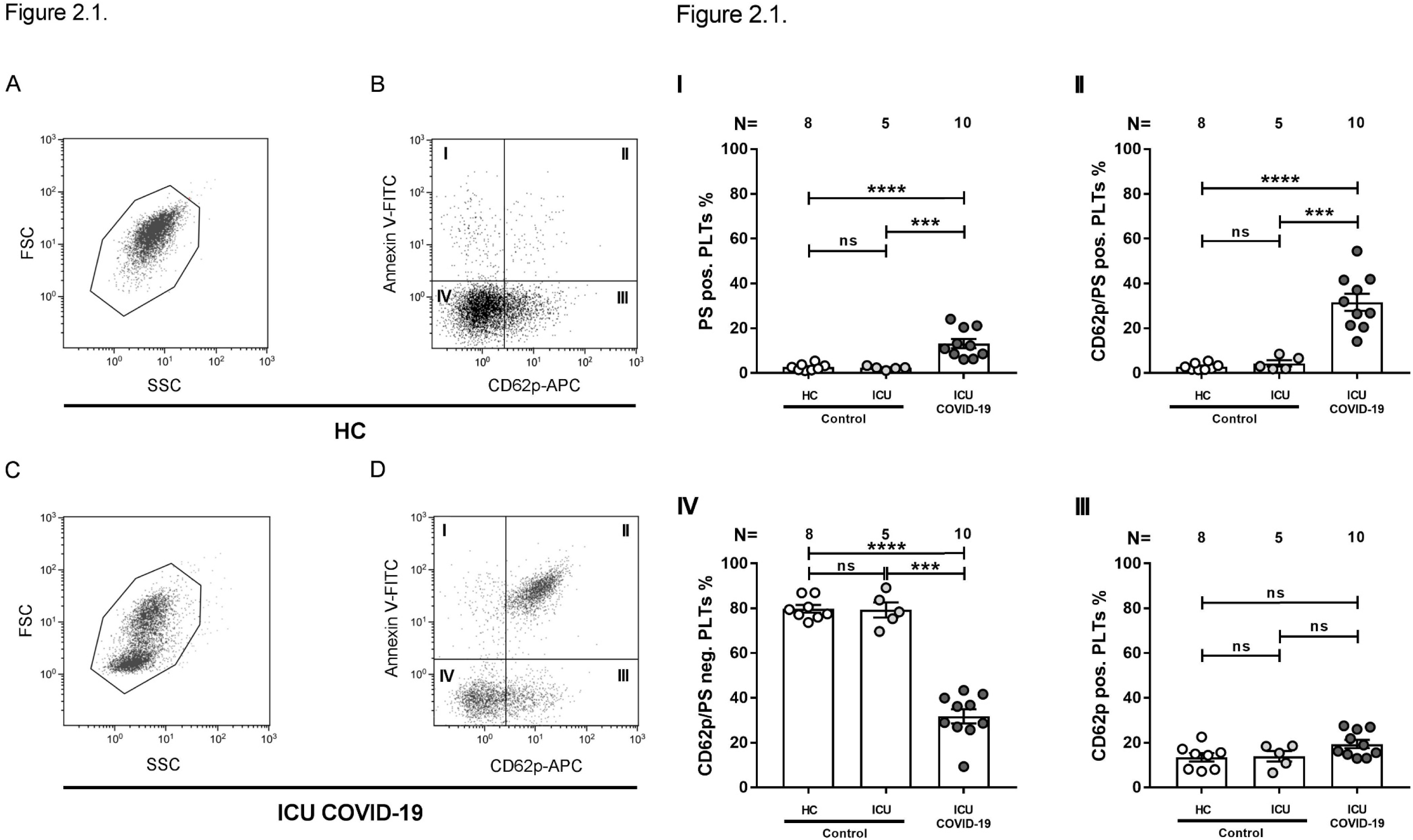
ICU COVID-19 IgGs induce procoagulant PLTs and increased clot formation. **Fig. 2.1**. (**A+C**) Representative FC plots of wPLTś FSC vs. SSC properties after HC or ICU COVID-19 IgG incubation. (**B+D**) Following HC or ICU COVID-19 IgG incubation, CD42a positive gated wPLTs were analyzed for PS externalization and expression of CD62p via Annexin V-FITC and CD62p-APC antibody staining, respectively. **Fig. 2.1**. (**I-IV**) represent quantitative gate distribution of CD42a positive events as indicated in **Fig. 2.1**. (**B+D**). Data are shown as percentage±SEM of Annexin V-FITC and or CD62p-APC positive labeled wPLTs after incubation with HC (n=8), ICU non-COVID-19 (n=5) or ICU COVID-19 IgG (n=10). ns, not significant; *p<0.05, **p<0.01, ***p<0.001 and ****p<0.0001. The number of patients and healthy donors tested is reported in each graph. HC, healthy control; N, number of HCs or patients; PS, phosphatidylserine.

Next, we sought to investigate the ability of IgG fractions from patients with severe COVID-19 to cause increased clot formation. PLTs from healthy individuals were incubated with IgGs from ICU COVID-19 patients or ICU and HCs, added to autologous whole blood samples and finally perfused throw collagen covered microfluidic channels at a shear rate of 1500 s^-1^ (60 dyne). As shown in Fig. 3 A, IgG from severe COVID-19 patients caused increased clot formation. Overall, significantly higher surface area coverage by thrombus (SAC) was observed in the presence of ICU COVID-19 IgGs compared to ICU controls and HCs (mean % SAC±SEM: 13.95%±1.55 vs. 2.86±1.10, p value 0.0070; and vs. 2.70±0.83, p value 0.0002, respectively, Fig. 3 B). Together with the increased percentage of procoagulant PLTs (CD62p/PS positive) in response to ICU COVID-19 IgGs, these findings might provide a potential explanation for the increased thromboembolic events in severely affected COVID-19 patients.

**Fig. 3.**
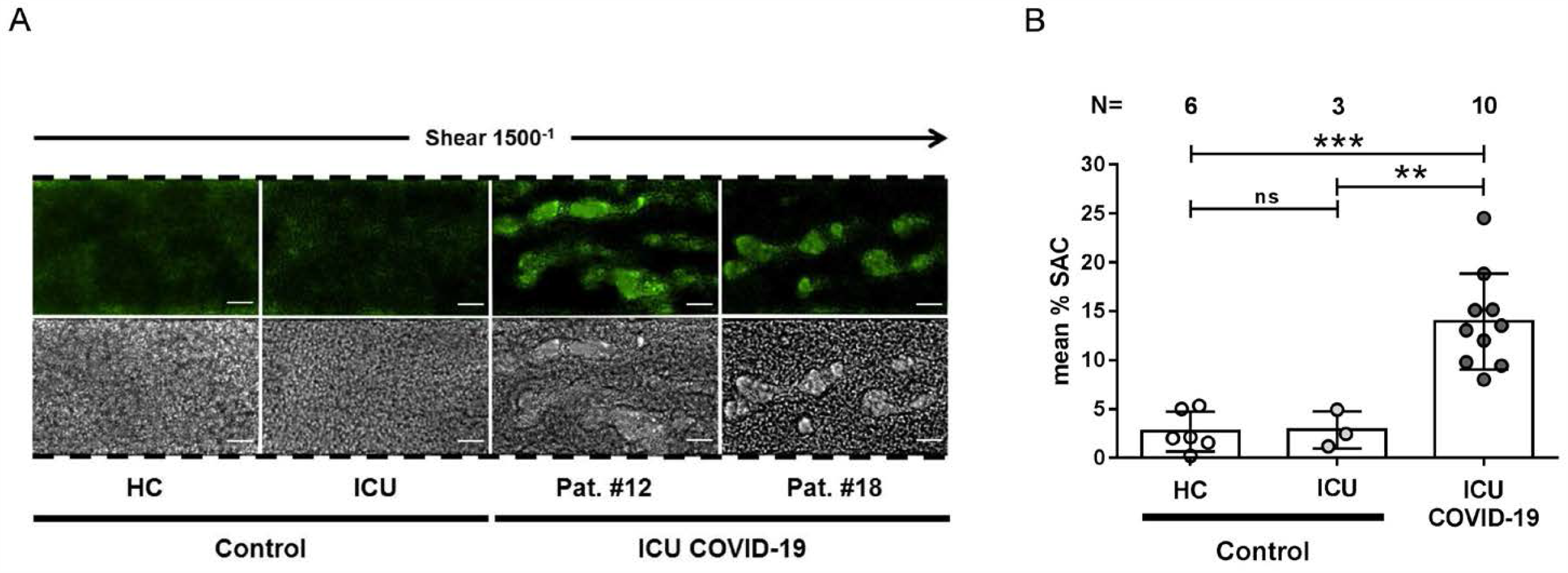
ICU COVID-19 IgG cause increased clot formation on collagen. (**A**) PRP from healthy individuals with the blood group O was incubated with HC (n=6), ICU non-COVID-19 control (n=3) or ICU COVID-19 IgG (n=10), labelled with FITC conjugated calcein and perfused through microfluidic channels at a shear rate of 1500^-1^ (60 dyne) for 5 min after reconstitution into autologous whole blood. Images were aquired at x20 magnification in the fluorescent (upper panel) as well as in the BF channel (lower panel). Scale bar 50µm. (**B**) Mean percentage of surface area covered (mean % SAC)±SEM by thrombus in the presence of HC (n=6), ICU non-COVID-19control (n=3) and ICU COVID-19 (n=10) IgG after 5 min perfusion time. ns, not significant; *p<0.05, **p<0.01, ***p<0.001 and ****p<0.0001. The number of patients and healthy donors tested is reported in each graph. HC, healthy control; N, number of HCs or patients.

### ICU COVID-19 IgGs cause procoagulant PLTs via crosslinking FcγRIIA

To further determine the underlying mechanistic pathways leading to ICU COVID-19 IgG induced formation of procoagulant PLTs, we next considered a potential ligation of FcγRIIA by patients, sera/IgGs. For this purpose, wPLTs were pretreated with the moAb IV.3. This FcγRIIA blockade resulted in marked inhibition of the antibody-induced Δψ depolarization (FI in % Δψ depolarization±SEM: 5.51±0.94 vs. 1.18±0.08, p value 0.0020, Fig. 4.1. A) as well as significant reduction of caspase activation (Ratio of cleaved caspase 3/procaspase 3±SEM: 4.70±1.16 vs. 1.33±0.29, p value 0.0286, Fig. 4.1. B+C). Intriguingly, the blockade of FcγRIIA almost abolished the changes in PLTś FSC and SSC properties (Fig. 4.2. A+C), and markedly reduced CD62p/PS double positive PLT population compared to isotype-control (% CD62p/PS positive PLTs±SEM: 48.91±3.05 vs. 12.88±1.65, p value 0.0078, Fig. 4.2. II).

**Fig. 4.**
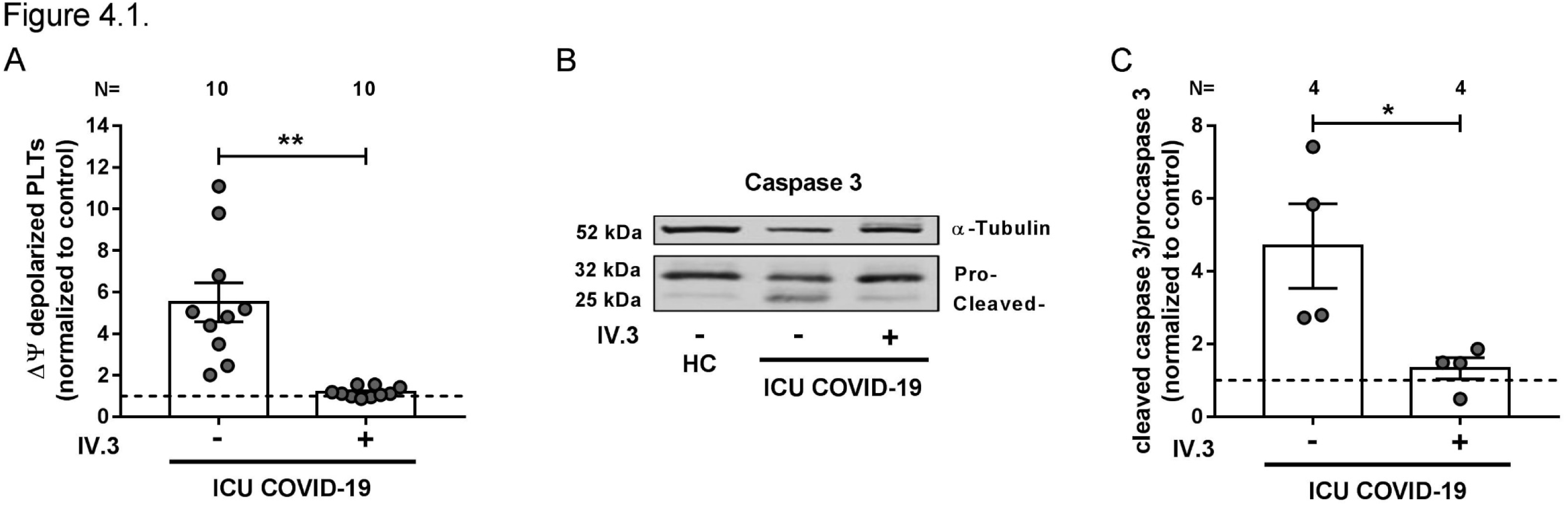
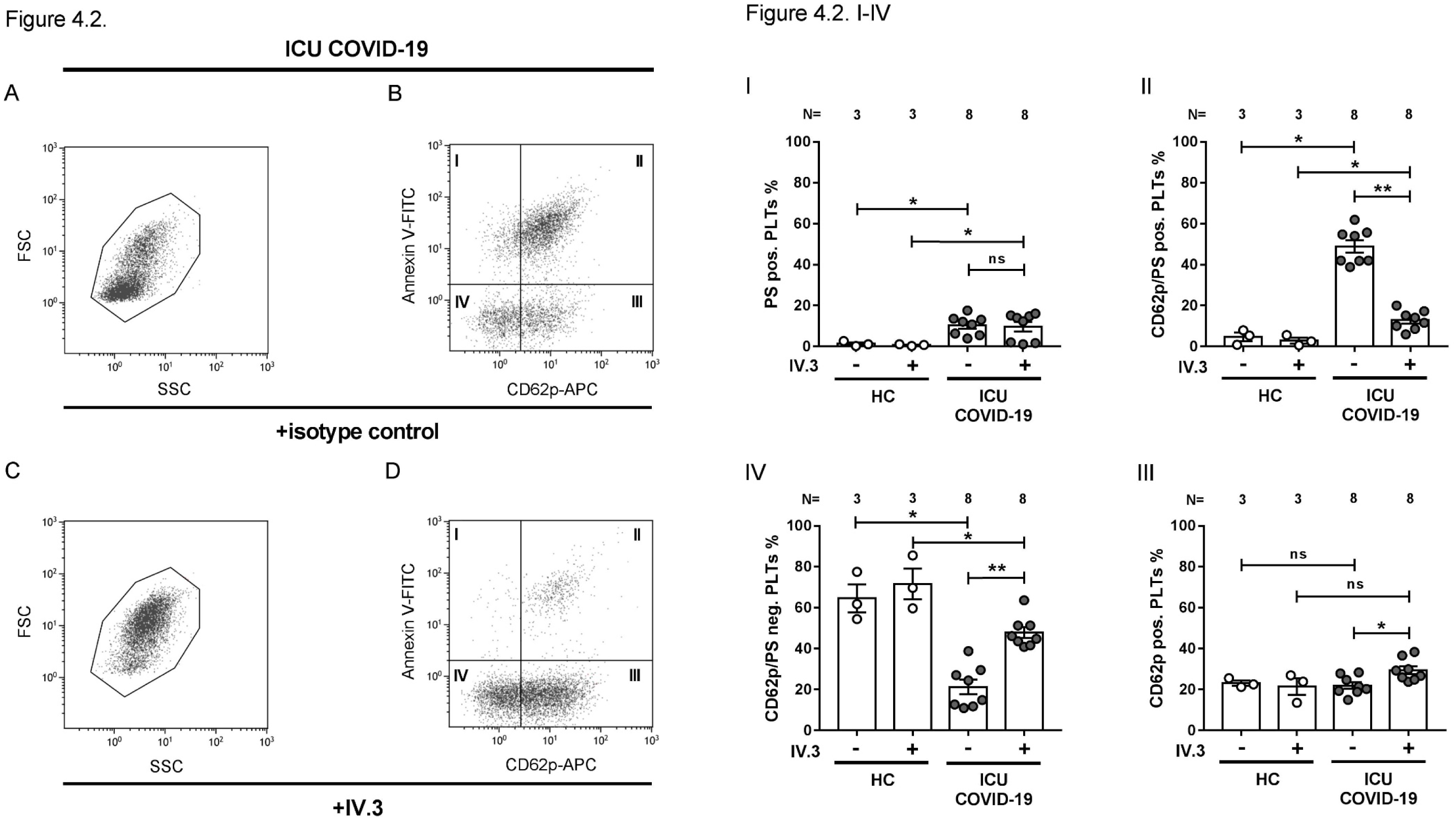
ICU COVID-19 IgG induced formation of procoagulant PLTs is FcγRIIA dependent. **Fig. 4.1. (A) ICU** COVID-19 patient serum (n=10) induced changes of Δψ in wPLTs were analyzed in the presence or absence of moAb IV.3 via FC. Data are presented as mean±SEM of the measured fold increase compared to control. (**B+C)** Representative WB image of detected cleaved caspase 3 (cleaved-) and procaspase 3 (pro-) levels in wPLTs that were incubated with ICU COVID-19 IgG (n=4) in the presence or absence of moAb IV.3. α-Tubulin served as loading control. (**C**) Densitometric analysis of cleaved caspase 3/procaspase 3 ratios from immunoblots as indicated in (**B**, n=4) normalized to control. **Fig. 4.2**. (**A+C**) Representative FC plots of wPLTś FSC vs. SSC after ICU COVID-19 IgG incubation in IV.3 pretreated wPLTs. (**B+D**) Gate distribution of CD42a positive PS (Annexin V-FITC) externalizing and CD62p (CD62p-APC) expressing wPLTs after ICU COVID-19 IgG incubation in isotype control or IV.3 pretreated wPLTs. **Fig. 4.2**. (**I-IV**) shows quantitative gate distributions of wPLTs with or without IV.3 pretreatment, after ICU COVID-19 IgG incubation as shown in **Fig. 4.2**. (**B+D**). Data are presented as percentage±SEM of Annexin V-FITC and or CD62p-APC positive labeled wPLTs after incubation with HC (n=3) or ICU COVID-19 IgG (n=8) in the presence or absence of moAb IV.3. ns, not significant; *p<0.05, **p<0.01, ***p<0.001 and ****p<0.0001. The number of patients and healthy donors tested is reported in each graph. HC, healthy control; N, number of HCs or patients; PS, phosphatidylserine.

Next, we analyzed the impact of FcγRIIA blockade on antibody-mediated clot formation. Pretreatment of PLTs with moAb IV.3 prior to ICU COVID-19 IgG incubation resulted in significant reduction of clot formation compared to isotype-control (mean % SAC±SEM: 16.49±1.02 vs. 5.84±1.93, respectively, p value 0.0090, Fig. 5 A+B). These results indicate that ICU COVID-19 IgG antibodies that are present in a subgroup of severe ICU COVID-19 patients have the capability to induce formation of procoagulant PLTs with increased clotting ability via crosslinking FcγRIIA.

**Fig. 5.**
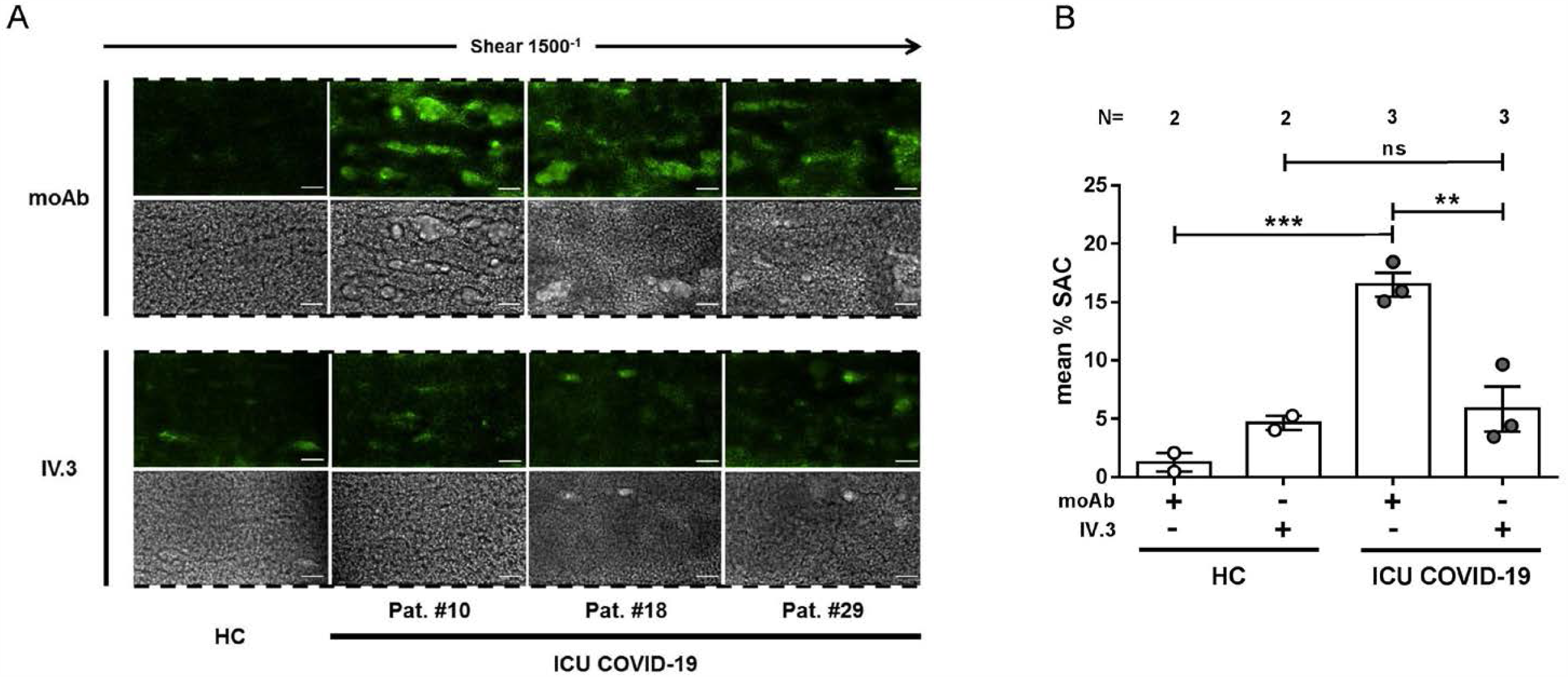
FcγRIIA inhibition prevents ICU COVID-19 IgG induced increased clot formation. **(A)** PRP from healthy individuals with the blood group O was incubated with HC (n=2) or ICU COVID-19 IgG (n=3 patients) in the presence of moAb IV.3 or isotype control (moAb) and perfused through microfluidic channels at a shear rate of 1500^-1^ (60 dyne) for 5 min. Images were aquired at x20 magnification in fluorescent (upper panel) as well as in the BF channel (lower panel). Scale bar 50µm. (**B**) mean % SAC±SEM induced by HC (n=2) or ICU COVID-19 IgG (n=3) in the presence or absence of moAb IV.3 or isotype control (moAb). ns, not significant; *p<0.05, **p<0.01, ***p<0.001 and ****p<0.0001. The number of patients and healthy donors tested is reported in each graph. moAb, monoclonal isotype control; HC, healthy control; N, number of HCs or patients

### Calcium is pivotal for the generation of ICU COVID-19-IgG induced procoagulant PLTs

Following the detection of an ICU COVID-19 IgG-induced procoagulant PLT phenotype, we sought to investigate the underlying intracellular molecular mechanisms. The contribution of calcium was analyzed by the depletion of extra- and intracellular calcium contents via EGTA and BAPTA, respectively. Extracellular calcium depletion caused significant inhibition of Δψ depolarization (FI in % Δψ depolarization±SEM: 3.08±0.18 vs. 1.94±0.20, p value 0.0079, Fig. 6.1. A) as well as marked reduction of caspase 3 cleavage (Ratio of cleaved caspase 3/procaspase 3±SEM: 4.14±0.65 vs. 1.50±0.20, p value 0.0079, Fig. 6.1. B+C). Moreover, depletion of extracellular calcium significantly inhibited ICU COVID-19 IgG induced alterations of wPLTś morphology (Fig. 6.2. A-D) as well as the generation of CD62p/PS positive PLTs (% CD62p/PS positive PLTs±SEM: 32.89±2.77 vs. 6.42±1.21, p value 0.0039, Fig. 6.2. II).

**Fig. 6.**
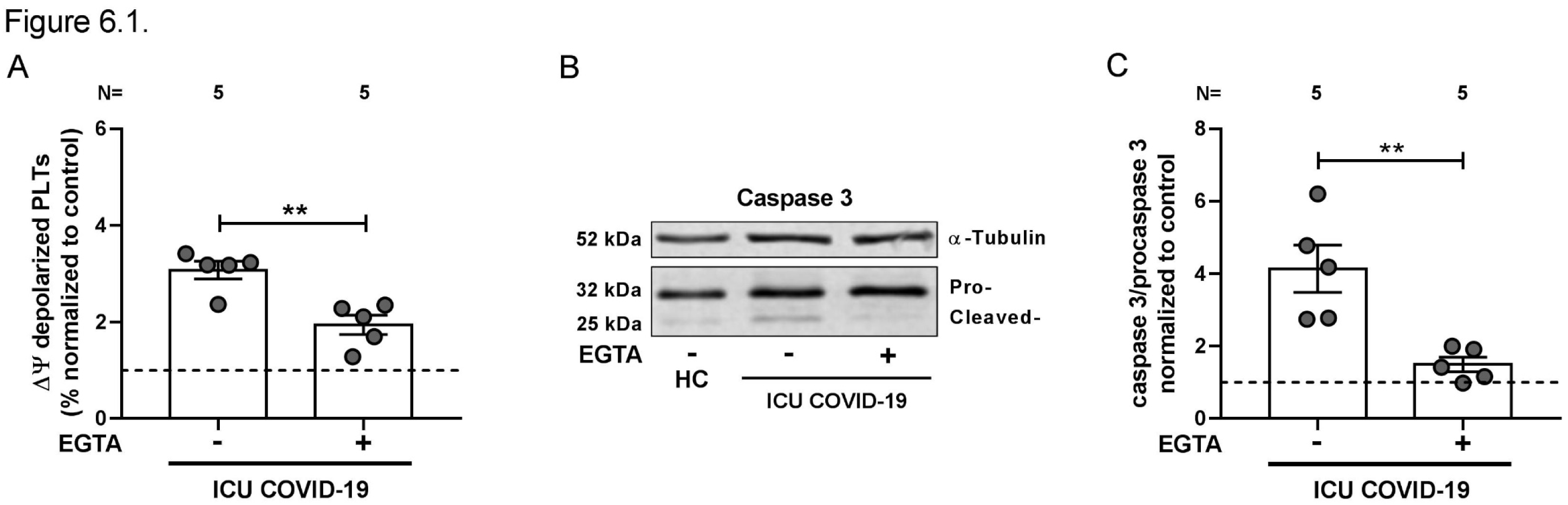
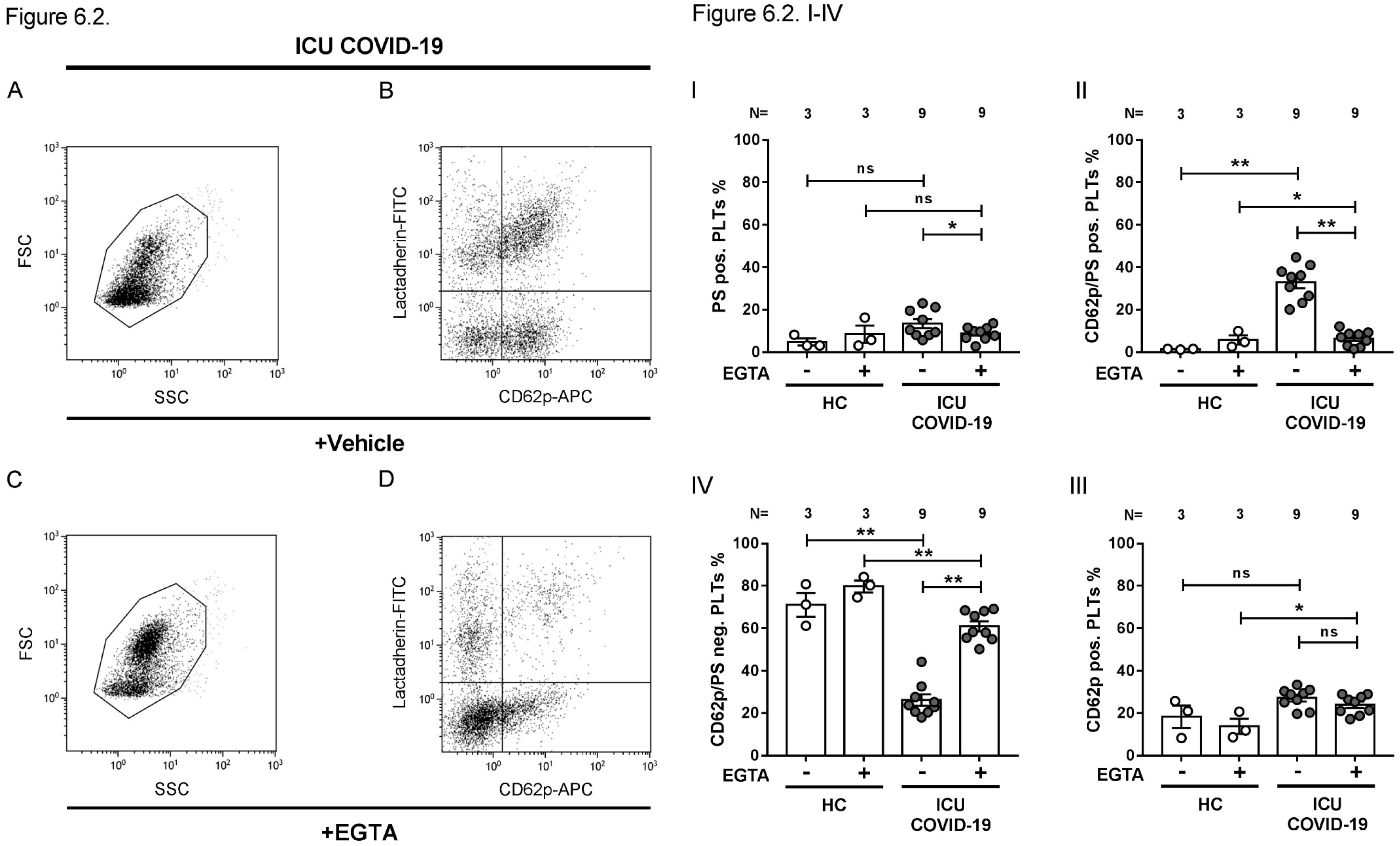
ICU COVID-19 IgG induced formation of procoagulant PLTs is dependent on extracellular calcium. **Fig. 6.1. (A)** FC assessment of ICU COVID-19 IgG (n=5) induced changes of Δψ in wPLTs that were pretreated with EGTA (1 mM) or vehicle. Data are presented as mean±SEM of the measured fold increase compared to control. (**B+C)** Representative WB image of detected cleaved caspase 3 (cleaved-) and procaspase 3 (pro-) levels in EGTA (1 mM) vehicle pretreated wPLTs that were incubated with ICU COVID-19 IgG (n=6), respectively. α-Tubulin served as loading control. (**C**) Densitometric analysis of immunoblots indicated in (**B**) for cleaved caspase 3/procaspase 3 ratios from WB data as indicated in (**B**, [n=5]) normalized to control. **Fig. 6.2**. (**A+C**) Representative FC plots of wPLTś FSC vs. SSC after ICU COVID-19 IgG incubation in EGTA (1mM) or vehicle pretreated wPLTs. (**B+D**) Gate distribution of CD42a positive PS (Annexin V-FITC) externalizing and CD62p (CD62p-APC) expressing wPLTs after ICU COVID-19 IgG incubation in EGTA (1mM) or vehicle pretreated wPLTs. **Fig. 6.2**. (**B+D**) Gate distribution of CD42a positive PS (Lactadherin-FITC) externalizing and CD62p (CD62p-APC) expressing wPLTs after ICU COVID-19 IgG incubation in EGTA (1 mM) or vehicle containing wPLTs. **Fig. 6.2**. (**I-IV**) shows quantitative gate distribution of wPLTs after incubation with ICU COVID-19 IgG. Data are presented as percentage±SEM of Lactadherin-FITC or CD62p-APC positive labeled wPLTs after incubation with HC (n=3) or ICU COVID-19 IgG (n=9) in vehicle or EGTA (1 mM) pretreated wPLTs. ns, not significant; *p<0.05, **p<0.01, ***p<0.001 and ****p<0.0001. The number of patients and healthy donors tested is reported in each graph. HC, healthy control; N, number of HCs or patients; PS, phosphatidylserine.

Similar effects were observed as depleting intracellular calcium stores. BAPTA treatment resulted in significant inhibition of ICU COVID-19 IgG-induced Δψ depolarization (FI in % Δψ depolarization±SEM: 4.46±0.73 vs. 0.99±0.09, p value 0.0039, Fig. 7.1. A) and caspase 3 cleavage (Ratio of cleaved caspase 3/procaspase 3±SEM: 3.50±0.98 vs. 0.43±0.06, p value 0.0286, respectively, Fig. 7.1. B+C). In addition, BAPTA pretreatment of wPLTs resulted in marked prevention of ICU COVID-19 IgG-induced changes in FSC and SSC (Figure 7.2. A-D as well as significant reduction of CD62p/PS positive PLTs (% CD62p/PS positive PLTs: 29.58±3.36 vs. 1.74±0.39, p value 0.0020, Fig. 7.2. II).

**Fig. 7.**
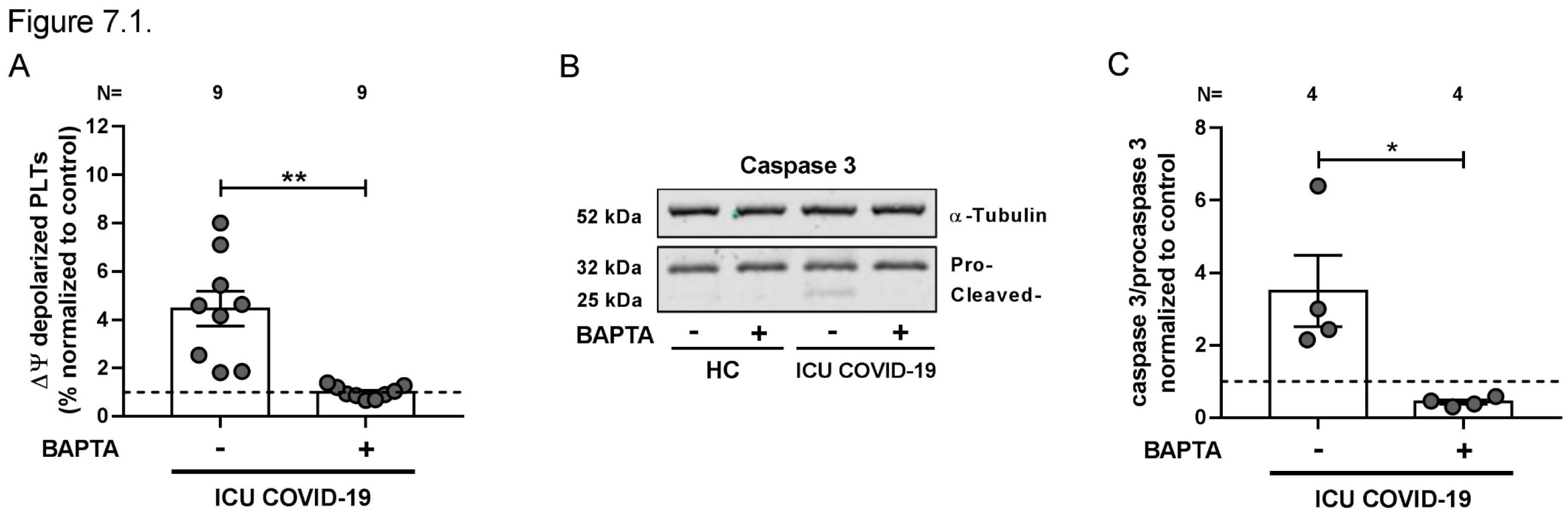
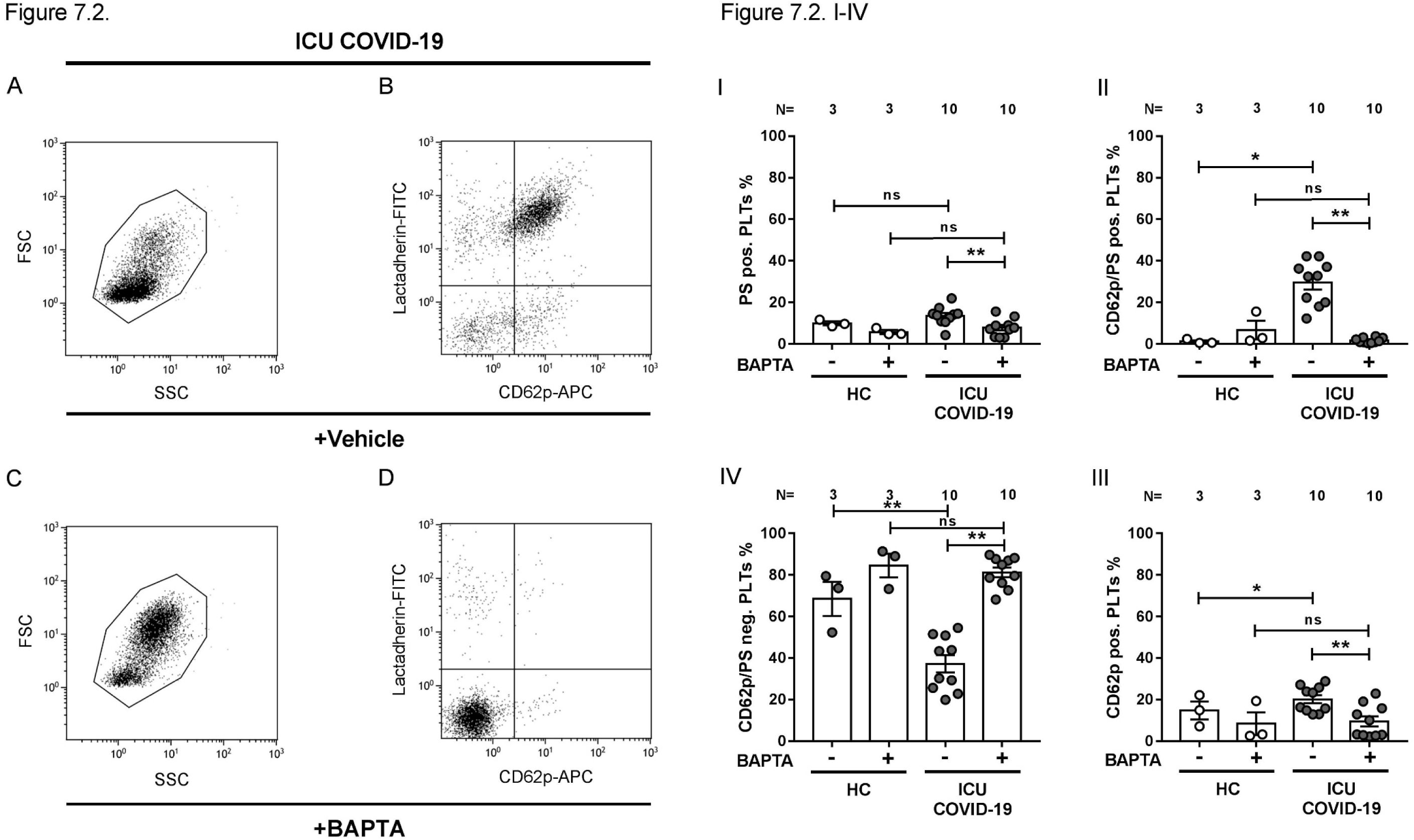
Depletion of intracellular calcium abrogates ICU COVID-19 IgG induced procoagulant PLT formation. **Fig. 7.1 (A) ICU** COVID-19 IgG (n=9) induced changes of Δψ in wPLTs were analyzed in the presence or absence of the intracellular calcium chelator BAPTA (20 µM). (**B)** Representative WB image of cleaved caspase 3 (cleaved-) and procaspase 3 (pro-) were detected with anti-caspase 3 antibody in wPLTs that were incubated with ICU COVID-19 IgG in BAPTA (20 µM) or vehicle pretreated wPLTs. α.Tubulin served as loading control. (**C**) Densitometric analysis of cleaved caspase 3/procaspase 3 ratios from immunoblots (indicated in **B**, [n=4]) normalized to control. **Fig. 7.2**. (**A+C**) FC plots of wPLTs FSC and. SSC after ICU COVID-19 IgG incubation in vehicle or BAPTA (20 µM) preloaded wPLTs. (**B+D**) FC plots of CD42a positive gated wPLTs that were incubated with ICU COVID-19 IgG in vehicle or in BAPTA (20 µM) preloaded wPLTs. **Fig. 7.2.(I-IV)** quantitative gate distribution of CD42a positive wPLTs based on the gate settings shown in **Fig. 7.2**. (**B+D**). Data are shown as percentage±SEM of Lactadherin-FITC and or CD62p-APC positive labeled wPLTs after HC (n=3) or ICU COVID-19 IgG (n=10) incubation in normal or BAPTA (20 µM) preloaded wPLTs. Ns,not significant; *p<0.05, **p<0.01, ***p<0.001 and ****p<0.0001. The number of patients and healthy donors tested is reported in each graph. HC, healthy control; N, number of HCs or patients, PS; phosphatidylserine.

### Activation of cAMP protects against ICU COVID-19 IgG induced procoagulant PLTs

The interplay between the signalling pathways of the intracellular second messengers, cAMP and calcium, has been shown to have an important role in numerous essential physiological processes during PLT activation and apoptosis (16, 17). Therefore, we investigated the role of cAMP on COVID-19 antibody-induced formation of procoagulant PLTs. Forskolin, an activator of intracellular ADC that elevates intracellular levels of cAMP, was recently reported to inhibit the formation of apoptotic PLTs in immune thrombocytopenia (18). The pretreatment of PLTs with Forskolin led to significant reduction of ICU COVID-19 IgG-induced Δψ depolarization (FI in % Δψ depolarization±SEM:5.10±0.70 vs. 1.43±0.17, p value 0.0079, Fig. 8.1. A) and caspase 3 cleavage (Ratio of cleaved caspase 3/procaspase 3±SEM: 3.74±1.14 vs. 1.01±0.14, p value 0.0313, respectively, Fig. 8.1. B+C). In addition, Forskolin pretreatment of wPLTs led to reduction of ICU COVID-19 IgG-induced changes in FSC and SSC properties (Fig. 8.2. A-D) as well as to a significant inhibition of procoagulant PLT generation (% CD62p/PS positive PLTs±SEM: 35.29±1.58 vs. 12.26±2.74, p value 0.0313, Fig. 8.2. II). These findings indicate that the elevation of intracellular cAMP prevents ICU COVID-19 IgG-induced formation of procoagulant PLTs.

**Fig. 8.**
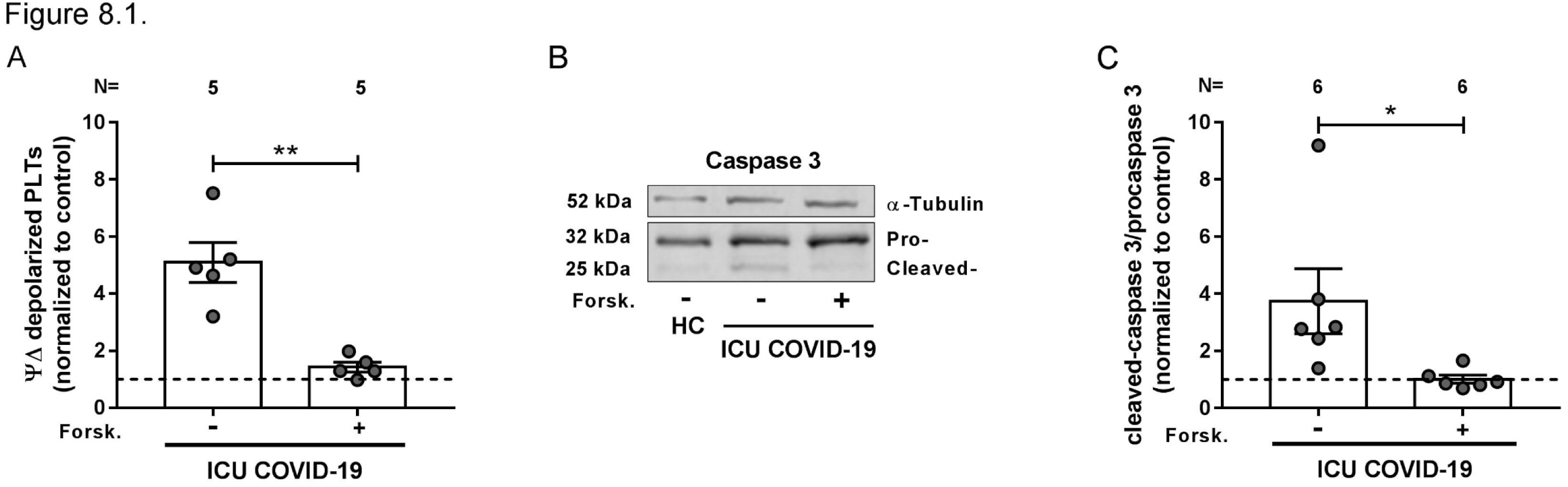
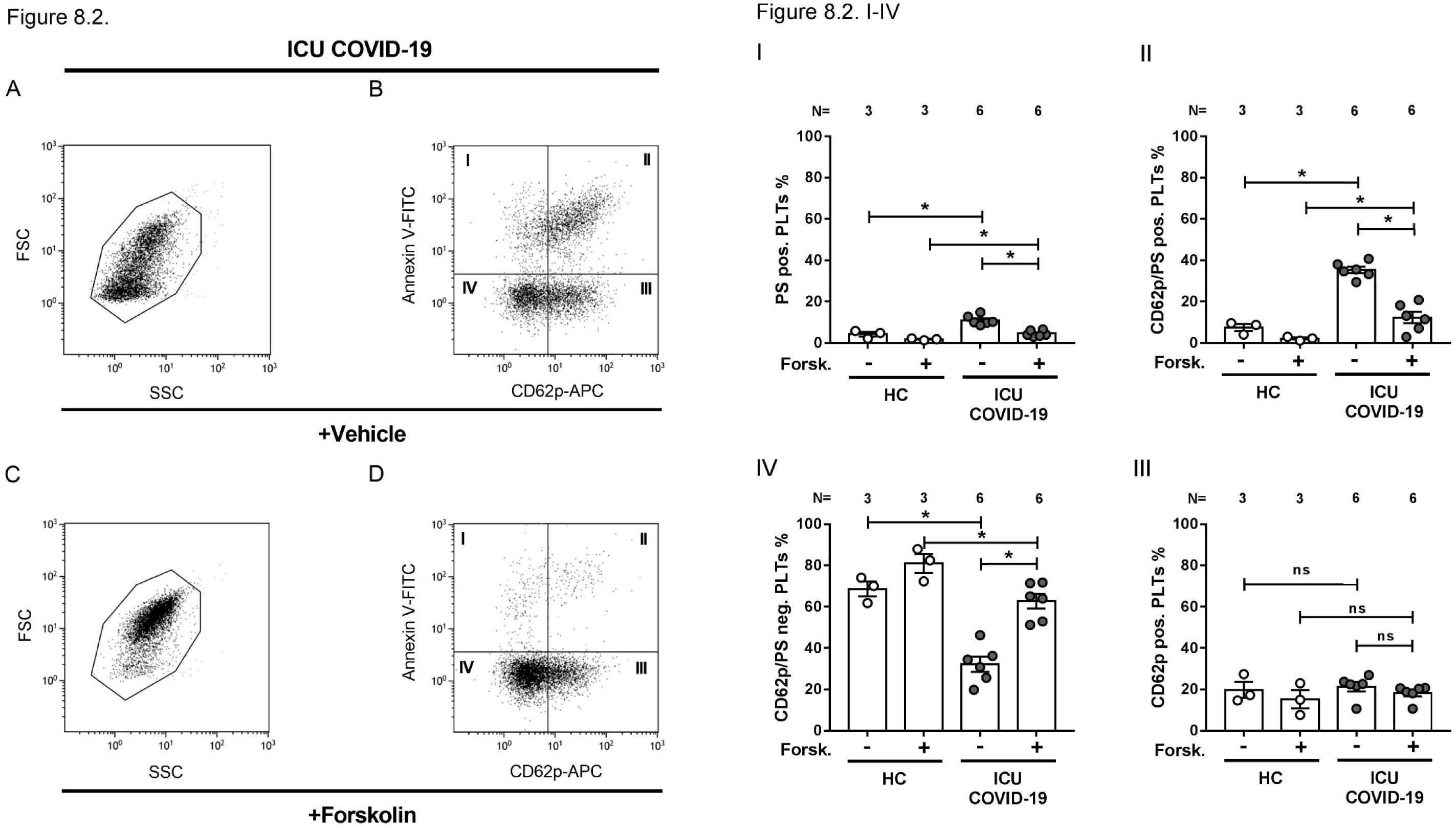
Forskolin protects ICU COVID-19 IgG induced procoagulant PLT formation. **Fig. 8.1. (A**) FC analysis of ICU COVID-19 IgG (n=5 patients) induced changes in Δψ of wPLTs, that were pretreated with vehicle or the ADC inductor Forskolin (2.25 µM) for 30 min at 37°C. Data are presented as mean±SEM of the measured fold increase compared to control. (**B**) Representative WB image of cleaved caspase 3 (cleaved-) and procaspase 3 (pro-) levels detected in wPLTs that were incubated with ICU COVID-19 IgG in vehicle or Forskolin (2.25 µM) pretreated wPLTs. α-Tubulin served as loading control. (**C**) Densitometric assesed ratios of cleaved caspase 3/procaspase 3 (as indicated in **B** [n=6]) normalized to control. **Fig. 8.2**. (**A+C**) FC plots of wPLTś FSC vs. SSC after ICU COVID-19 IgG incubation in vehicle or Forskolin (2.25 µM) pretreted wPLTs. (**B+D**) Gate distribution of CD42a positive wPLTs that were incubated with ICU COVID-19 IgG vehicle or Forskolin (2.25 µM) pretreated wPLTs. **Fig. 8.2. (I-IV)** Quantitative distribution of CD42a positive wPLTs based on the gate settings shown in **Fig. 8.2**. (**B+D**). Data are shown as percentage ±SEM of Annexin V-FITC and or CD62p-APC positive labeled wPLTs that were incubated with IgGs from HC (n=3) or ICU COVID-19 IgG (n=6) in vehicle or Forskolin (2.25 µM) pretreated wPLTs. ns, not significant; *p<0.05, **p<0.01, ***p<0.001 and ****p<0.0001. The number of patients and healthy donors tested is reported in each graph. HC, healthy control; N, number of HCs or patients; PS, phosphatidylserine.

More importantly and of high clinical interest, similar protective effect was observed with Iloprost, an already approved cAMP inducer (19). In fact, Iloprost pretreatment of wPLTs led to marked reductions of ICU COVID-19 IgG-induced Δψ depolarization (FI of % Δψ depolarization±SEM: 4.66±0.57 vs. 1.77 ±0.32, p value 0.0078, Fig. 9.1. A), and cleavage of caspase 3 (Ratio of cleaved caspase 3/procaspase 3±SEM: 4.69±1.45 vs. 2.0348±0.38, p value 0.0156, respectively, Fig. 9.1. B+C). In addition, Iloprost pretreatment led to a significant reduction of changes in PLT morphology (Fig. 9.2. A-D) as well as in the number of procoagulant CD62p/PS positive PLTs (% CD62p/PS positive PLTs±SEM: 41.36±3.60 vs. 22.22±3.92, p value 0.0156, Fig. 9.2. II). Noteworthy, no significant changes were observed in the number of CD62p single positive PLTs (% CD62p positive PLTs±SEM: 24.48±2.2 vs. 26.51±3.94, p value 0.6875).

**Fig. 9.**
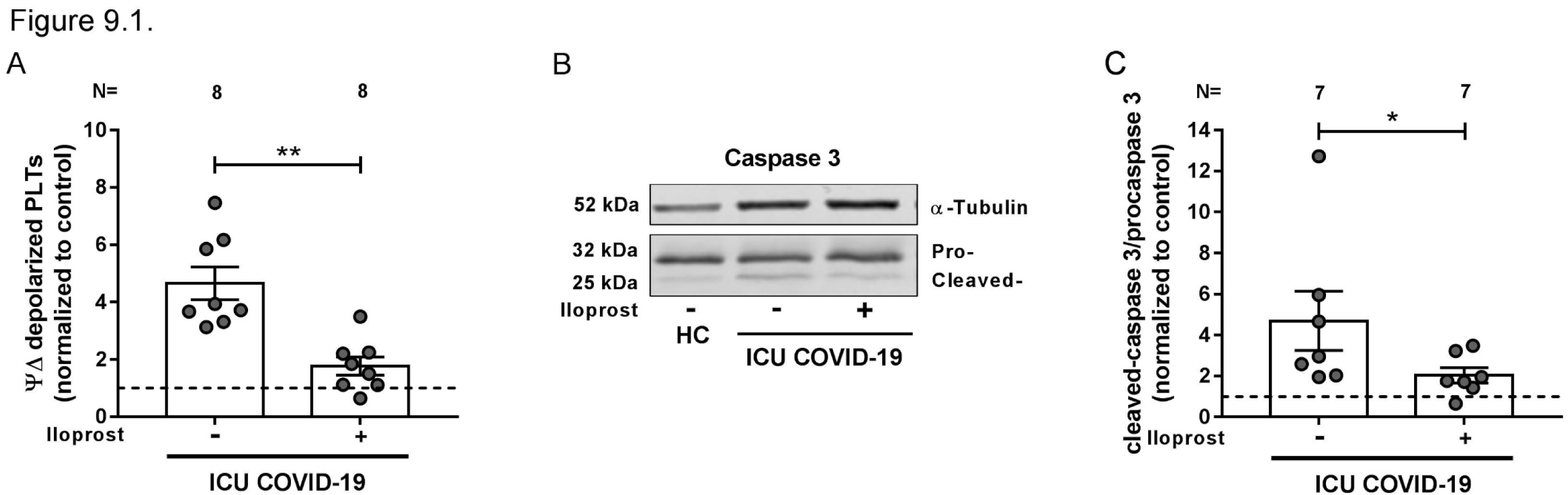
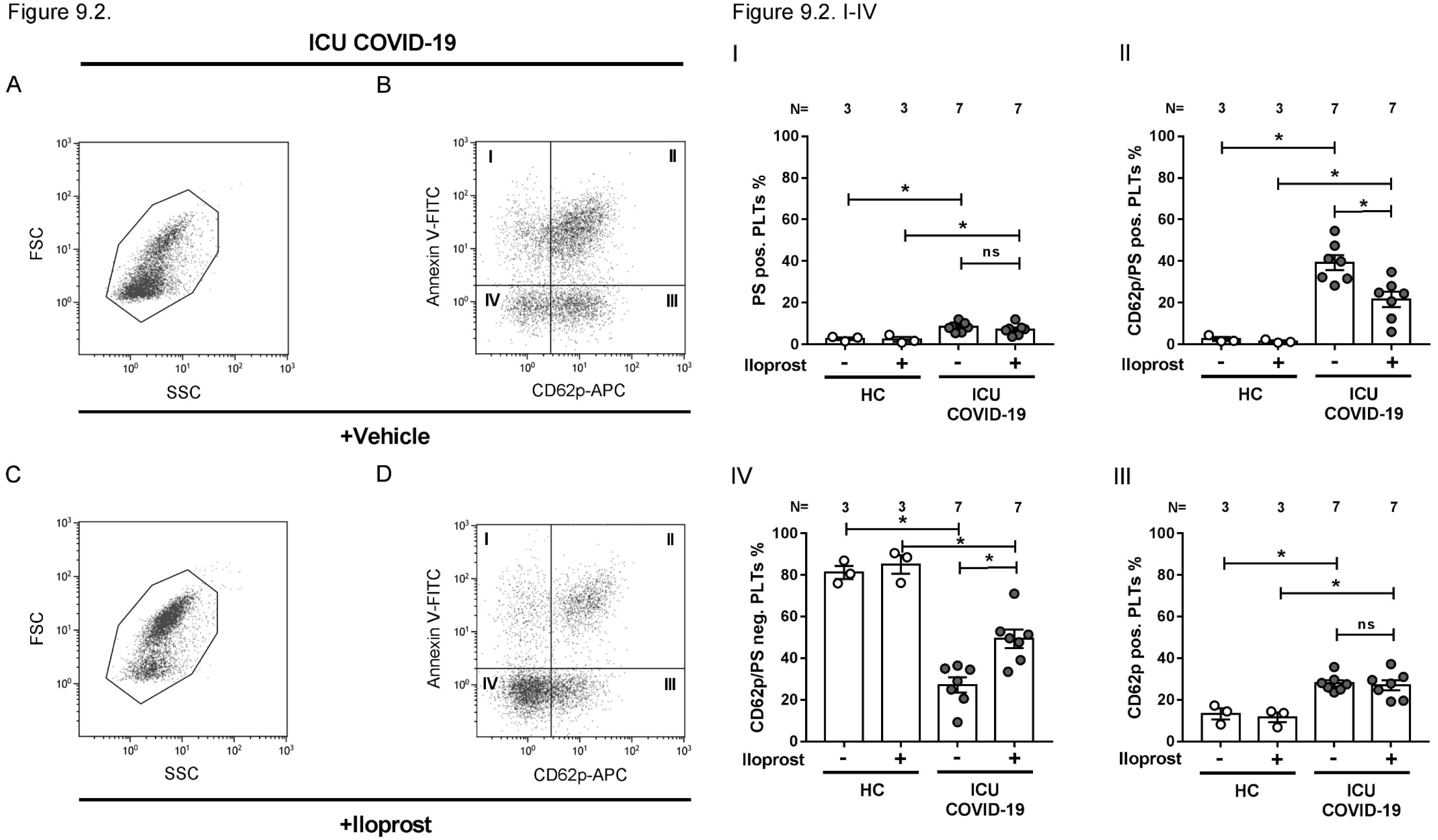
cAMP elevation via Iloprost prevents ICU COVID-19 IgG induced procoagulant PLTs. **Fig. 9.1.(A)** Changes in Δψ depolarization induced by ICU COVID-19 IgG (n=8 patients) were analyzed in vehicle or Iloprost (20 nM) pretreated wPLTs via FC. Data are presented as mean±SEM of the measured fold increase compared to control. **(B)** Representative WB image of detected caspase 3 (cleaved-) and procaspase 3 (pro-) levels in wPLTs that were pretreated with vehicle or Iloprost (20 nM) prior to ICU COVID-19 IgG incubation. α-Tubulin served as loading control. **(C)** Densitometric analysis of cleaved caspase 3/procaspase 3 ratios from the WB data (indicated in **B**, [n=7]) normalized to control. **Fig. 9.2. (A+C)** FC detected changes in wPLTś FSC vs. SSC properties after ICU COVID-19 IgG incubation in vehicle or Iloprost (20 nM) pretreated wPLTs. **Fig. 9.2**. (**B+D**) (**E+G**) Gate distribution of CD42a positive vehicle or Iloprost (20 nM) pretreated wPLTs that were incubated with ICU COVID-19 IgG. **Fig. 9.2. (I-IV)** Quantitative distribution of the CD42a positive wPLT population based on the gate settings shown in **Fig. 9.2**. (**B+D**). Data are shown as percentage±SEM of Annexin V-FITC and or CD62p-APC positive labeled wPLTs that were incubated with HC (n=3) or ICU COVID-19 IgG (n=6) in vehicle or Iloprost (20 nM) pretreated wPLTs. ns, not significant; *p<0.05, **p<0.01, ***p<0.001 and ****p<0.0001. The number of patients and healthy donors tested is reported in each graph. HC, healthy control; N, number of HCs or patients; PS, phosphatidylserine.

Based on these findings we were interested whether Iloprost might affect the ability to form blood clots. Pretreatment of PLTs with Iloprost showed a marked reduction in ICU COVID-19 IgG-induced clot formation compared to vehicle (mean % SAC±SEM: 14.63±2.31 vs. 3.85±0.95, respectively, p value 0.0079, Fig. 10. A+B). These findings provide the first evidence for a potential therapeutic use of Iloprost in the treatment of the antibody-induced coagulopathy that is observed in COVID-19 disease.

**Fig. 10.**
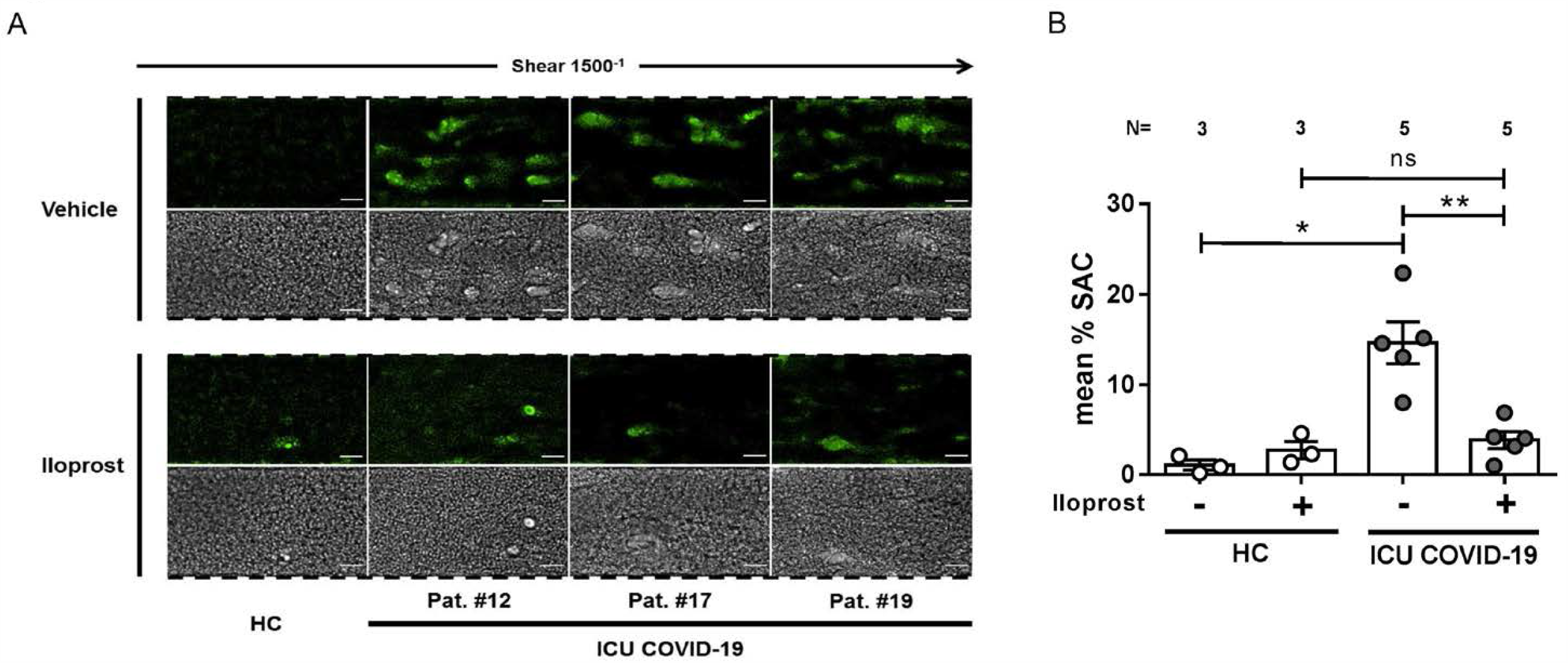
Iloprost inhibits ICU COVID-19 IgG induced increased clot formation. **(A)** PRP from healthy individuals with the blood group O was incubated with HC (n=3) or ICU COVID-19 IgG (n=5) in the presence of vehicle or Iloprost (20 nM). After resonstituion into autologous whole blood, samples were perfused through microfluidic channels at a shear rate of 1500^-1^ (60 dyne) for 5 min. Pictures were aquired at x20 magnification in fluorescent (upper panel) as well as in the BF channel (lower panel). Scale bar 50µm. (**B**) Mean % SAC±SEM induced by HC (n=3) or ICU COVID-19 IgG (n=5) in the presence of vehicle or Iloprost (20 nM). ns,not significant; *p<0.05, **p<0.01, ***p<0.001 and ****p<0.0001. The number of patients and healthy donors tested is reported in each graph. HC, healthy control; N, number of HCs or patients; PS, phosphatidylserine.

## Discussion

Our study showed that IgGs from patients with severe COVID-19 are able to induce procoagulant PLTs with increased potential for clot formation via crosslinking FcγRIIA in a calcium depending manner. Most importantly, we showed that cAMP activation by an approved drug, namely Iloprost, can sufficiently inhibit the initiation of procoagulant PLTs and the subsequent increased clot formation. These data may have tremendous clinical implications.

Meanwhile, it is well established that COVID-19 infection is associated with systemic prothrombotic state and increased incidence of thromboembolic complications (20). However, the pre-sequelae events leading to the coagulopathy observed in COVID-19 still remains elusive. Data from our study demonstrate the presence of PLT-reactive IgG antibodies that harbour the ability to induce marked changes in PLTs in terms of increased Δψ depolarization, PS externalization and P-selectin expression, which are characteristic for procoagulant PLTs. A novel finding, and potentially of high clinical interest, is that antibody-mediated formation of procoagulant PLTs was associated with the clinical course as these changes progressively increased as patients needed ventilation and peaked between day 3 and 7 on ICU. Probably, the kinetic of these markers might have predictive value to monitor COVID-19-induced coagulopathy. In fact, PS externalization was recently found to be a predictive biomarker for thromboembolic complications related to cardiovascular therapeutic devices (21). In contrast to PS, conventional markers of PLT activation were declared in this study to fail the detection of early activation events leading to thrombosis.

The alterations in PLTs that were observed in our study after incubation with sera from patients with severe COVID-19 infection, such as Δψ disruption, caspase 3 cleavage and PS externalization, could be found in apoptotic as well as procoagulant PLTs. The involvement of PLT apoptosis to promote prothrombotic conditions has been controversially discussed. In fact, recent data suggests that apoptotic PLTs are unable to promote prothrombotic conditions (10). However, a clear dissection of the molecular events leading to prothrombotic PLTs is challenging since activation of the apoptosis caspase pathway has been described in the late phase of agonist-induced PLT activation as well as in PLTs from patients with chronic kidney disease (22, 23), which are prone to thromboembolic events. Our findings showed that antibody-mediated Δψ disruption and caspase cleavage is associated with the induction of a CD62p/PS positive PLT population suggesting that IgG antibodies in COVID-19 induce procoagulant rather than apoptotic PLTs. In fact, activated PLTs were shown elsewhere to be predominant in COVID-19 patients (6). In particular, CD62p positive PLTs were suggested to be involved in thrombosis in COVID-19 via the interaction with neutrophil granulocytes leading to NET formation and increased interaction with the inflamed vessel wall. Our data showed that COVID-19 IgG-antibodies trigger a PLT population with procoagulant potential. An important finding that was reinforced by data from an ex vivo microfluidic circulation system which revealed an increased clot formation in the presence of COVID-19 IgG-antibodies.

Motivated by these novel functional data, we thought to dissect the exact mechanistic pathways involved in the COVID-19 IgG-induced procoagulant status. We found that ICU COVID-19 IgG-mediated changes in wPLTs involves the ligation of the immune receptor FcγRIIA. The blockade of FcγRIIA significantly inhibits ICU COVID-19 IgG-induced changes in Δψ depolarization as well as cleavage of caspase 3. Most importantly, FcγRIIA blockade reduced procoagulant CD62p/PS positive PLTs and subsequently inhibited COVID-19 antibody-induced ex vivo clot formation. While this findings are novel for COVID-19-associated coagulopathy, the correlation between FcγRIIA ligation and increased risk for thromboembolic events is well established for heparin-induced thrombocytopenia (HIT) (24). Interestingly, the IgG antibody formation peaks in HIT within 5-10 day after exposure to heparin and is associated with PLT consumption and increased risk for thrombosis (25). Similarly, in our study the ability of ICU COVID-19 sera to induce procoagulant PLTs was most pronounced between day 3 and 7. Of note, antibody-induced alterations in PLT markers were accompanied by enhanced levels of IgG antibodies against the Spike S epitope of SARS-CoV-2.These findings might suggest a transient onset of misdirected autoimmune mechanisms that result in the emergence of PLT-reactive antibodies leading to a prothrombotic status in severe COVID-19 infection.

Calcium is involved in many biological mechanistic pathways in PLTs (16). In response to PLT agonists, calcium is released from the PLT internal stores which is followed by amplifying second wave extracellular calcium influx via store (SOCE) and receptor operated calcium entry (ROCE), respectively (26). The role of calcium signalling in ITAM (immunoreceptor tyrosine-based activation motif) coupled GPVI and (hem)ITAM coupled CLEC-2 receptor signalling has been well established in the last few years (27). Additionally, FcγRIIA ligation has been shown to induce calcium mobilization via ITAM signalling prior to platelet aggregation(28). In our study, the depletion of calcium in the extracellular compartment inhibited ICU COVID-19 IgG-induced procoagulant changes. This finding indicates that alterations in SOCE or ROCE might be involved in the antibody-mediated generation of procoagulant PLTs in COVID-19. In fact, the regulative role of SOCE for procoagulant PS externalization has been previously reported, as SOCE channel inhibition resulted in reduced PS externalization in human erythroleukemia cells (29). In addition, reduced PS externalization and decreased clot formation was observed in chimeric mice that express mutated impaired SOCE calcium channel Orai1 R93W on the PLT surface, indicating that SOCE is a major inductor of PLT PS externalization and procoagulant PLT formation (30). A potential role of ITAM-regulated signalling leading to procoagulant PLTs has been also suggested. Calcium depletion inhibited GP VI induced formation of procoagulant PLTs (31). In this study, the authors proposed that distinct signalling cascades, most likely tyrosine and extracellular calcium dependent, could be involved in the formation of procoagulant PLTs. Our findings emphasize the role of extracellular calcium in FcγRIIA (ITAM) mediated procoagulant PLT formation. Another possible explanation for these finding could be the loss of distinct calcium-dependent conformational properties of PLT epitopes that are targeted by COVID-19 IgG antibodies. Dimeric PLT GP IIb/IIIa, the receptor for fibrinogen and well known immunogenic epitopes of PLT-reactive autoantibodies in immune thrombocytopenia (ITP), has been well characterized to be structurally dependent on extracellular calcium levels (32). Although this might be currently too speculative, the depletion of extracellular calcium might have inhibited antibody binding to such conformation-sensitive epitopes on GP IIb/IIIa. ICU COVID-19 IgG-induced PLT alterations were dependent on extracellular as well as intracellular calcium levels. Since PLT apoptosis has been described to be independent of calcium (33), our finding indicates that COVID-19 IgGs trigger FcγRIIA-mediated events that result in procoagulant PLT formation in a calcium-dependent manner. In fact, similar findings were reported by recently showing that actin-mediated PLT shape change and phosphoinositide 3-kinase activity in response to FcγRIIA ligation in dependent on intracellular calcium (28). The absence of ICU COVID-19 IgG-mediated PLT changes could be also due to inactivation of calcium dependent cysteine protease calpain that is crucial for the conformational changes in GP IIb/IIIa (34, 35). Another possible explanation could be the inactivation of calcium dependent TMEM16F that bears essential properties for membrane phospholipid scrambling and microparticle generation (10). Future studies could explore the exact mechanisms by which calcium contributes to procoagulant PLT formation in COVID-19.

COVID-19 antibody-induced procoagulant PLTs were significantly inhibited by the use of inducers of adenylate cyclase (ADC) that are well known to cause increased cAMP levels in PLTs (36). The protective effect of cAMP was further demonstrated, as Iloprost, an already approved prostacyclin derivate and ADC, efficiently prevented the formation of procoagulant PLTs in response to COVID-19 antibodies. Finally and of high clinical importance, Iloprost pretreatment of PLTs markedly reduced COVID-19 IgG-induced clot formation on collagen suggesting that cAMP inducers may have potential to prevent life threatening thromboembolic events in COVID-19 antibody-mediated coagulopathy. Another minor finding from our microfluidic system was that Iloprost, although significantly inhibited antibody-mediated thrombus formation, did not affect the CD62p-single positive population. Since Iloprost prevented clot formation, this finding might indicate that PS rather than CD62p exposure on the PLT surface is pivotal to trigger the onset of thromboembolic events.

Our study is subjected to some limitations. First, as an observational, monocentric study, we cannot conclude that the reported associations between IgG antibodies and changes in activation/apoptosis markers in COVID-19 are causal for the thrombosis or specific for the disease. Second, we cannot exclude the possibility of remaining residual confounding or unmeasured potential confounders in our mechanistic studies. Third, the low number of patients does not enable a final and robust statistical analysis to assess clinical outcomes in patients with increased procoagulant PLTs. Nevertheless, data presented in this study may provide a background for future studies to dissect mechanisms related to PLT activation that are involved in the progression of COVID-19.

Taken together, our study shows that IgG antibodies from patients with severe COVID-19 are able to stimulate FcγRIIA leading to the induction of procoagulant PLTs with an increased ability of clot formation. These processes are dependent on calcium and can be efficiently inhibited by cAMP inducers suggesting that ADC might represent a potentially promising target to prevent thromboembolic complications in COVID-19 disease.

## Data Availability

Data are available on request to the corresponding author

## Author contributions

J.Z., K.A., T.B. and P.R. designed the study. P.R. and H.H. were responsible for the treatment of the patients. K.A. and H.H. collected and analyzed the clinical data. J.Z., K.A., H.J., L.P., A.S. and A.W. performed the experiments. J.Z., K.A., H.J., L.P., A.S. and A.W. collected the data. J.Z., K.A., H.J., L.P., A.S., A.W., V.M., P.R. and T.B. analyzed the data, interpreted the results and wrote the manuscript. All authors read and approved the manuscript.

## Acknowledgements

This work was supported by grants from the “Ministerium für Wissenschaft, Forschung und Kunst Baden-Württemberg” to J.Z. and T.B. and from the Herzstiftung to T.B. (BA5158/4 and TSG-Study) and TÜFF-Gleichstellungsförderung to K.A. (2563-0-0). We thank Karoline Weich for her excellent technical support. The authors thank Susanne Staub for editing the article as native English speaker.

## References

1. Zhou F, Yu T, Du R, Fan G, Liu Y, Liu Z, et al. Clinical course and risk factors for mortality of adult inpatients with COVID-19 in Wuhan, China: a retrospective cohort study. Lancet. 2020;395(10229):1054–62.

2. Hanff TC, Mohareb AM, Giri J, Cohen JB, Chirinos JA. Thrombosis in COVID-19. Am J Hematol. 2020;95(12):1578–89.

3. Iba T, Levy JH, Levi M, Connors JM, Thachil J. Coagulopathy of Coronavirus Disease 2019. Crit Care Med. 2020;48(9):1358–64.

4. Henry BM, Vikse J, Benoit S, Favaloro EJ, Lippi G. Hyperinflammation and derangement of renin-angiotensin-aldosterone system in COVID-19: A novel hypothesis for clinically suspected hypercoagulopathy and microvascular immunothrombosis. Clin Chim Acta. 2020;507:167–73.

5. Hottz ED, Azevedo-Quintanilha IG, Palhinha L, Teixeira L, Barreto EA, Pao CRR, et al. Platelet activation and platelet-monocyte aggregate formation trigger tissue factor expression in patients with severe COVID-19. Blood. 2020;136(11):1330–41.

6. Manne BK, Denorme F, Middleton EA, Portier I, Rowley JW, Stubben C, et al. Platelet gene expression and function in patients with COVID-19. Blood. 2020;136(11):1317–29.

7. de Witt SM, Verdoold R, Cosemans JM, Heemskerk JW. Insights into platelet-based control of coagulation. Thromb Res. 2014;133 Suppl 2:S139–48.

8. Swieringa F, Spronk HMH, Heemskerk JWM, van der Meijden PEJ. Integrating platelet and coagulation activation in fibrin clot formation. Res Pract Thromb Haemost. 2018;2(3):450–60.

9. Agbani EO, Poole AW. Procoagulant platelets: generation, function, and therapeutic targeting in thrombosis. Blood. 2017;130(20):2171–9.

10. Reddy EC, Rand ML. Procoagulant Phosphatidylserine-Exposing Platelets in vitro and in vivo. Front Cardiovasc Med. 2020;7:15.

11. Hua VM, Chen VM. Procoagulant platelets and the pathways leading to cell death. Semin Thromb Hemost. 2015;41(4):405–12.

12. Greinacher A, Michels I, Kiefel V, Mueller-Eckhardt C. A rapid and sensitive test for diagnosing heparin-associated thrombocytopenia. Thromb Haemost. 1991;66(6):734–6.

13. Marini I, Zlamal J, Faul C, Holzer U, Hammer S, Pelzl L, et al. Autoantibody-mediated desialylation impairs human thrombopoiesis and platelet life span. Haematologica. 2019.

14. Mangin PH, Gardiner EE, Nesbitt WS, Kerrigan SW, Korin N, Lam WA, et al. In vitro flow based systems to study platelet function and thrombus formation: Recommendations for standardization: Communication from the SSC on Biorheology of the ISTH. J Thromb Haemost. 2020;18(3):748–52.

15. Huang C, Wang Y, Li X, Ren L, Zhao J, Hu Y, et al. Clinical features of patients infected with 2019 novel coronavirus in Wuhan, China. Lancet. 2020;395(10223):497–506.

16. Mammadova-Bach E, Nagy M, Heemskerk JWM, Nieswandt B, Braun A. Store-operated calcium entry in thrombosis and thrombo-inflammation. Cell Calcium. 2019;77:39–48.

17. Nagy Z, Smolenski A. Cyclic nucleotide-dependent inhibitory signaling interweaves with activating pathways to determine platelet responses. Res Pract Thromb Haemost. 2018;2(3):558–71.

18. Zhao L, Liu J, He C, Yan R, Zhou K, Cui Q, et al. Protein kinase A determines platelet life span and survival by regulating apoptosis. J Clin Invest. 2017;127(12):4338–51.

19. Fisch A, Michael-Hepp J, Meyer J, Darius H. Synergistic interaction of adenylate cyclase activators and nitric oxide donor SIN-1 on platelet cyclic AMP. Eur J Pharmacol. 1995;289(3):455–61.

20. Tang N, Li D, Wang X, Sun Z. Abnormal coagulation parameters are associated with poor prognosis in patients with novel coronavirus pneumonia. J Thromb Haemost. 2020;18(4):844–7.

21. Roka-Moiia Y, Walk R, Palomares DE, Ammann KR, Dimasi A, Italiano JE, et al. Platelet Activation via Shear Stress Exposure Induces a Differing Pattern of Biomarkers of Activation versus Biochemical Agonists. Thromb Haemost. 2020;120(5):776–92.

22. Shcherbina A, Remold-O’Donnell E. Role of caspase in a subset of human platelet activation responses. Blood. 1999;93(12):4222–31.

23. Bonomini M, Dottori S, Amoroso L, Arduini A, Sirolli V. Increased platelet phosphatidylserine exposure and caspase activation in chronic uremia. J Thromb Haemost. 2004;2(8):1275–81.

24. Chong BH, Fawaz I, Chesterman CN, Berndt MC. Heparin-induced thrombocytopenia: mechanism of interaction of the heparin-dependent antibody with platelets. Br J Haematol. 1989;73(2):235–40.

25. Warkentin TE. Clinical picture of heparin-induced thrombocytopenia (HIT) and its differentiation from non-HIT thrombocytopenia. Thromb Haemost. 2016;116(5):813–22.

26. Rink TJ, Sage SO. Calcium signaling in human platelets. Annu Rev Physiol. 1990;52:431–49.

27. Rayes J, Watson SP, Nieswandt B. Functional significance of the platelet immune receptors GPVI and CLEC-2. J Clin Invest. 2019;129(1):12–23.

28. Barkalow KL, Falet H, Italiano JE, Jr., van Vugt A, Carpenter CL, Schreiber AD, et al. Role for phosphoinositide 3-kinase in Fc gamma RIIA-induced platelet shape change. Am J Physiol Cell Physiol. 2003;285(4):C797–805.

29. Kunzelmann-Marche C, Freyssinet JM, Martinez MC. Regulation of phosphatidylserine transbilayer redistribution by store-operated Ca2+ entry: role of actin cytoskeleton. J Biol Chem. 2001;276(7):5134–9.

30. Bergmeier W, Oh-Hora M, McCarl CA, Roden RC, Bray PF, Feske S. R93W mutation in Orai1 causes impaired calcium influx in platelets. Blood. 2009;113(3):675–8.

31. Heemskerk JW, Vuist WM, Feijge MA, Reutelingsperger CP, Lindhout T. Collagen but not fibrinogen surfaces induce bleb formation, exposure of phosphatidylserine, and procoagulant activity of adherent platelets: evidence for regulation by protein tyrosine kinase-dependent Ca2+ responses. Blood. 1997;90(7):2615–25.

32. Tomiyama Y, Kosugi S. Autoantigenic epitopes on platelet glycoproteins. Int J Hematol. 2005;81(2):100–5.

33. Schoenwaelder SM, Yuan Y, Josefsson EC, White MJ, Yao Y, Mason KD, et al. Two distinct pathways regulate platelet phosphatidylserine exposure and procoagulant function. Blood. 2009;114(3):663–6.

34. Mattheij NJ, Gilio K, van Kruchten R, Jobe SM, Wieschhaus AJ, Chishti AH, et al. Dual mechanism of integrin alphaIIbbeta3 closure in procoagulant platelets. J Biol Chem. 2013;288(19):13325–36.

35. Kulkarni S, Jackson SP. Platelet factor XIII and calpain negatively regulate integrin alphaIIbbeta3 adhesive function and thrombus growth. J Biol Chem. 2004;279(29):30697–706.

36. Smolenski A. Novel roles of cAMP/cGMP-dependent signaling in platelets. J Thromb Haemost. 2012;10(2):167–76.

