## Supplementary material for "cAMP prevents antibody-mediated thrombus formation in COVID-19": Suppl.

### Supplementary Fig. 1

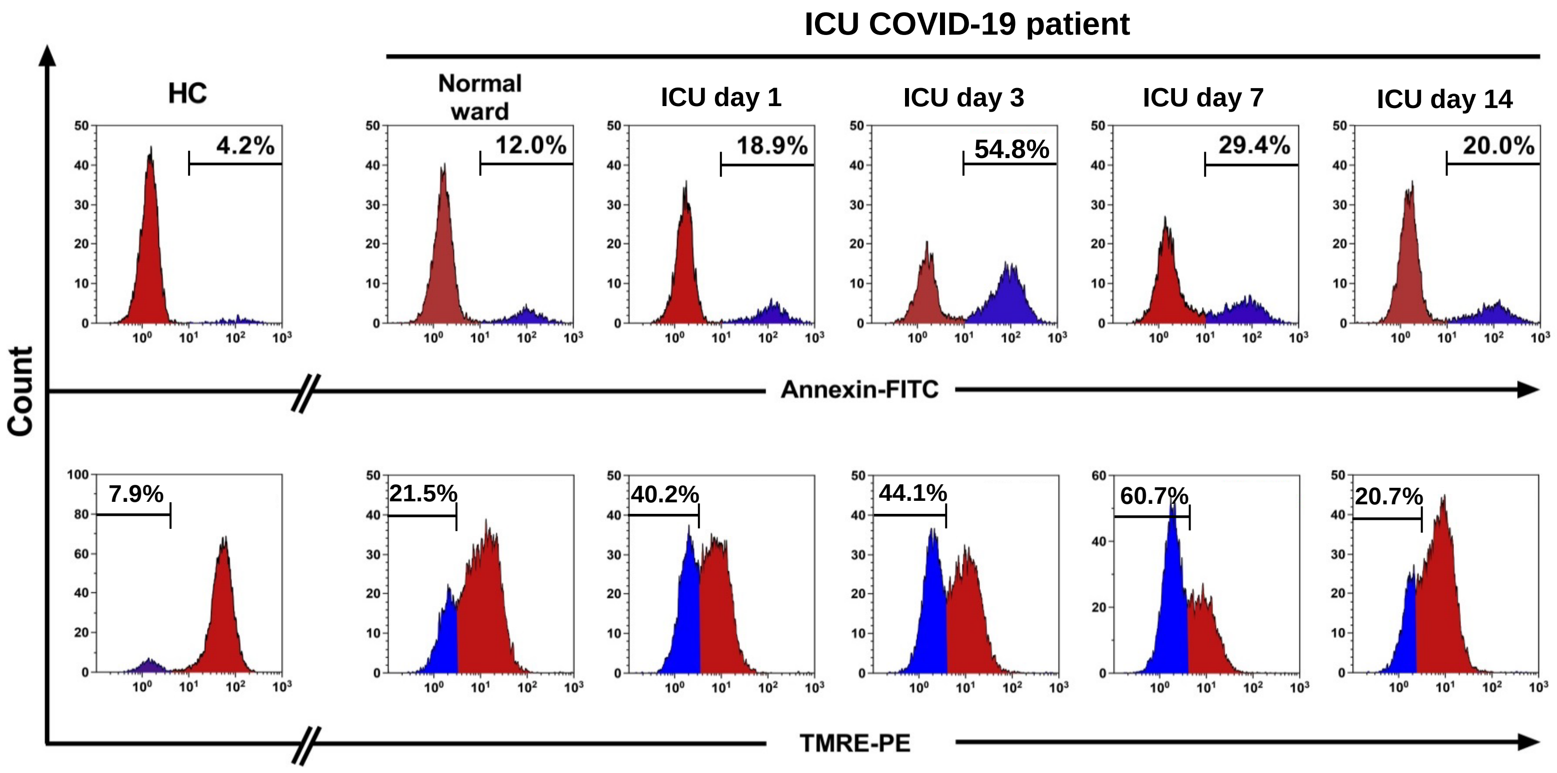


**Supplementary Fig. 1-FC detected changes of PLT PS externalization and Δψ depolarization during severe COVID-19 disease.** Histograms represent the kinetics of changes in PS externalization (upper panel) as well as Δψ depolarization (lower panel) induced by sera of one COVID-19 patient. Serum samples were withdrawn at hospital admission with a moderate state of COVID-19 disease (normal ward) as well as during the patients´ severe course of disease at day 1, 3, 7 and 14 of ICU hospitalization. COVID-19 serum induced changes in PLT PS externalization as well as Δψ depolarization were detected by FC using Annexin V-FITC and TMRE-PE staining, respectively. Data are presented as percentage of Annexin positive and TMRE negative events, respectively. HC, healthy control.

### Supplementary Fig. 2 A

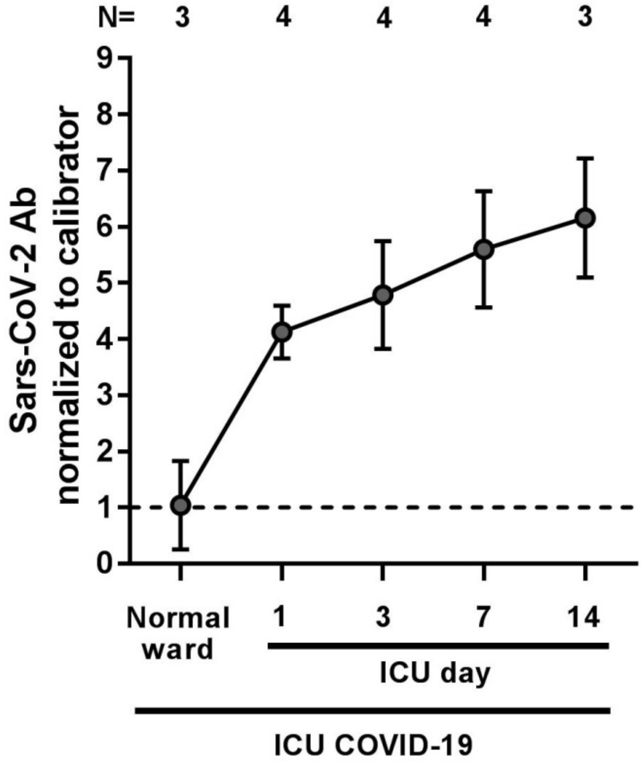


**Supplementary Fig. 2 A- SARS-CoV-2 antibody titres of COVID-19 patients during severe course of disease patients.** Follow up sera of ICU COVID-19 patients were withdrawn with moderate (n=3) as well as with severe COVID-19 disease at day 1, 3, 7 and 14 of ICU hospitalization (n=4) and screened for SARS-CoV-2 IgG-antibodies against the S1 spike protein by ELISA testing. SARS-CoV-2 IgG antibody titres were evaluated semi-quantitatively by calculation of a ratio of the extinction of the control or patient sample over the extinction of the calibrator. The cut-off for samples to be considered positive was a ratio ≥0.8. Dot lines represent baseline of the calibrator. The number of patients and healthy donors tested is reported in each graph.

### Supplementary Fig. 2 B

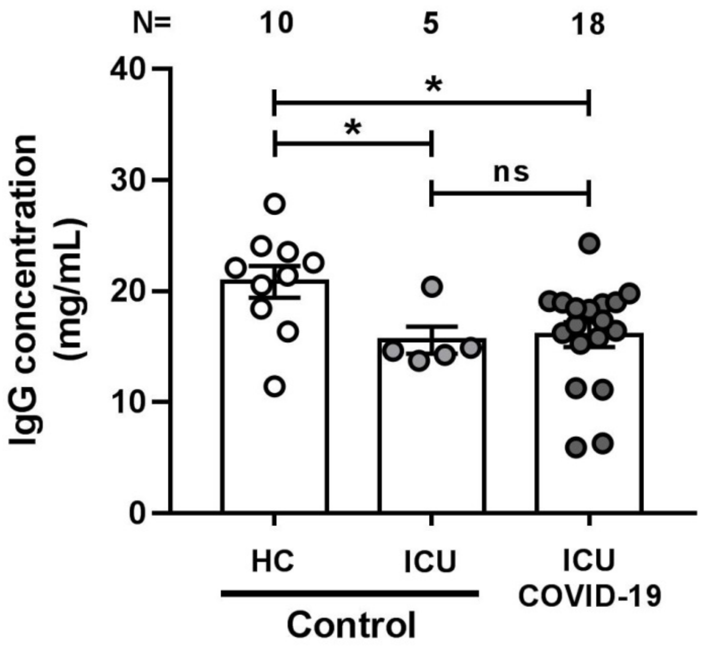


**Supplementary Fig. 2 B- Concentrations of IgG fractions isolated from HC-, ICU non-COVID-19 control and ICU COVID-19 patient sera.** After isolation from serum, concentrations of HC, ICU non-COVID-19 control or ICU COVID-19 IgG fractions were assessed using a NanoDrop One spectrophotometer. ns, not significant; *$p<0.05$, **$p<0.01$, ***$p<0.001$ and ****$p<0.0001$. The number of patients and healthy donors tested is reported in each graph.

### Supplementary Fig. 3

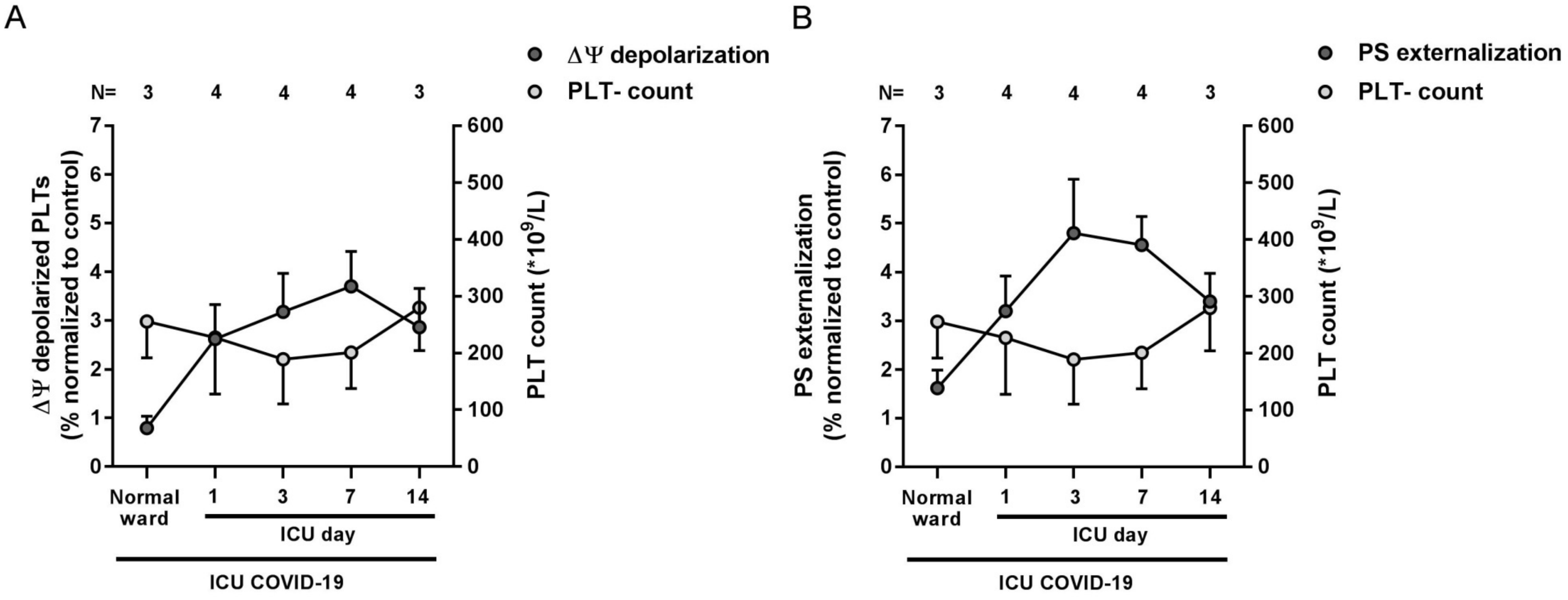


**Supplementary Fig. 3-Serum induced changes of PS externalization and Δψ depolarization vs. PLT-counts.** Graphs show serum induced changes in Δψ (**A**) as well as PS externalization (**B**) in wPLTs that were induced by sera of 4 COVID-19 patients that were collected for up to 14 days during hospitalization. In analogue to serum induced changes, PLT-counts of the corresponding patients on the day of serum sample withdrawal are shown (grey circles). Data are presented as mean±SEM of the measured fold increase compared to control (black circles) or absolute PLT-counts. The number of patient sera tested is reported in each graph. Dot lines in (**A+B**) represent the calculated cutoffs determined testing sera from healthy donors as mean of fold increase (FI) + 2xSEM. The number of patients and healthy donors tested is reported in each graph.
